# Predictors of COVID-19 vaccine acceptance across time and countries

**DOI:** 10.1101/2020.12.09.20246439

**Authors:** John R. Kerr, Claudia R. Schneider, Gabriel Recchia, Sarah Dryhurst, Ullrika Sahlin, Carole Dufouil, Pierre Arwidson, Alexandra L. J. Freeman, Sander van der Linden

## Abstract

Understanding the drivers of vaccine acceptance is crucial to the success of COVID-19 mass vaccination campaigns. Across 25 national samples from 12 different countries we examined the psychological correlates of willingness to receive a COVID-19 vaccine (total N = 25,334), with a focus on risk perception and trust in a number of relevant actors, both in general and specifically regarding the COVID-19 pandemic. Male sex, trust in medical and scientific experts and worry about the virus emerge as the most consistent predictors of reported vaccine acceptance across countries. In a subset of samples we show that these effects are robust after controlling for attitudes towards vaccination in general. Our results indicate that the burden of trust largely rests on the shoulders of the scientific and medical community, with implications for how future COVID-19 vaccination information should be communicated to maximize uptake.

## Introduction

COVID-19 has resulted in over a million deaths globally, illness for millions more, and unprecedented social and economic disruption^1,2^. Many governments have signaled that mass vaccination against the virus is the most straightforward—and possibly only—route to normality and stability^3,4^. While recent announcements of effective vaccines^5,6^ are promising, the wider impact of vaccines on preventing the spread of disease is also dependent on the uptake within a given population. In order to achieve ‘herd immunity’, enough people in a population must be immune to prevent the spread of a disease among non-immune individuals. The proportion varies depending on a number of factors including how infectious the contagion is, its prevalence in a population, and the variation in individual susceptibility or exposure to infection^7^. Estimates for the level of immunity required for COVID-19 herd immunity have ranged from 50% to 80% of the population, acquired through either natural infection and recovery, or through vaccination^8,9^.

Vaccine hesitancy—defined as a delay in acceptance or refusal of vaccines despite availability^10^—poses a challenge to achieving herd immunity. If a sufficient number of people in a population reject vaccination—and herd immunity is not achieved—the virus will continue to circulate among susceptible individuals, including those who are unable to be vaccinated for medical reasons. The WHO identified vaccine hesitancy as one of the top 10 threats to global health in 2019^11^, and in the pressing context of COVID-19, understanding vaccine hesitancy has only grown in importance^12^.

Public health researchers concerned with uptake of vaccination have understandably sought to uncover the drivers of vaccine hesitancy. By identifying antecedents of vaccine hesitancy, policy makers, public health officials, and professional communicators can target interventions to increase uptake of vaccines and ultimately reduce the burden of disease in a population^4^. However, strategies developed for campaigns targeting diseases with well-established vaccines (e.g. MMR, pertussis) may not fully translate to a pandemic context where there is greater uncertainty, less information available, and where institutional trust plays a greater role—as was noted in the wake of the 2009 H1N1 influenza pandemic^13^.

Recent evidence shows that acceptance of a COVID-19 vaccine is far from universal in many countries. Lazarus et al^14^ conducted a series of surveys across 19 countries in June 2020, asking respondents how much they agreed with the following statement: ‘If a COVID-19 vaccine is proven safe and effective and is available, I will take it’. The proportion of respondents who agreed ranged from 88.6% (China) to 55.8% (Russia). Examining possible predictors of vaccine acceptance, the authors report that men, older people, and those who express greater trust in the government were more likely to express willingness to receive a vaccine. The role of trust (in science, the government or the medical system) is a recurring theme in many other recent studies which have examined COVID-19 vaccine hesitancy in individual countries^15–20^. For example, Palamenghi et al^20^ report that across two large random samples of the Italian population, trust in science was positively correlated (*r* = .37) with willingness to receive a COVID-19 vaccine. Frank and Arim^16^ report that Canadians who are more trusting of local and national government bodies are more likely to express intentions to receive a vaccine if available, as are those who report high general social trust (i.e. believing that ‘most people can be trusted’).

Such results align with pre-COVID studies which have highlighted the role of trust in vaccination intentions and attitudes^13,21,22^. However, we note that recent studies examining COVID-19 vaccine intentions have typically only examined trust in one entity (e.g. government or hospitals); research to date has not considered the possible overlap between trust in the government, trust in science and medicine, and general social trust^23–25^. There is also a question over the extent to which vaccine acceptance is linked to mistrust in experts and authorities *regarding COVID-19 in particular*, or a more general lack of trust in these actors. In order to target communications specifically designed to satisfy the information needs of those who distrust official authorities, it is important to identify the precise agents that they distrust (and, ideally, why).

Beyond trust, the perceived threat or risk posed by a given disease has also been shown to predict vaccination attitudes. Models of health behavior, such as the Health Belief Model^26^ and Protection Motivation Theory^27^, place the perceived risk or severity of a disease as a key driver of vaccination intentions (and other preventative health behaviors)^13,28^. Recent surveys in the US, Malaysia, and Israel have shown that perceived risk and worry regarding the COVID-19 virus is associated with vaccine acceptance^29–31^. Other factors, such as the perceived benefits and costs as well as efficacy of protective behaviors are also outlined in models of health behavior as predictors of engagement in a given health behavior. However, until recently, little information about the possible costs, distribution and efficacy of a COVID-19 vaccine was available, meaning that the public has not generally been able to assess the potential benefits of a vaccine outside of a purely hypothetical arena (although experimental work has examined the influence of these factors on willingness to receive a vaccine^32^).

There are also increasing concerns about the politicization of science and about politics becoming entangled with vaccine beliefs and attitudes specifically, particularly in the context of a pandemic where central government structures are deeply involved in all stages of the public health response^13,33^. Prior research^34^ has shown that the rhetoric adopted by political elites on social media can fuel anti-vaccination attitudes amongst their followers and that ideologies can help explain anti-vaccination attitudes^30,35^.

In the current study we present a more comprehensive international analysis of the role of key social, political, and psychological predictors of COVID-19 vaccine acceptance across 12 countries, with multiple national surveys in some countries (total *N* = 25,334, see Table 1). All samples were recruited via online panel providers using quotas to ensure samples were matched to the general population in terms of age and gender (with the exception of France, see methods). Unlike previous studies, we examine reported trust in a range of actors, both in general and specifically relating to the COVID-19 pandemic. We also include several demographic factors (including politics), numeracy (known to play a role in risk perceptions^36^, and vaccine attitudes in particular^37^), affective (worry) and cognitive (perceived likelihood of infection) aspects of perceived COVID-19 risk^38^, broad measures of perceived efficacy, and, in a subset of samples, general attitudes towards vaccines.

**Table 1.** Sample demographics and percentage of participants willing to receive a COVID-19 vaccine or to recommend it to vulnerable friends/family.

| Country | Source | Date | N | M<sub>Age</sub> (SD) | Female (%) | Tertiary Educated (%) | Vaccine - acceptance (%) | Vaccine - recommend (%) |
| --- | --- | --- | --- | --- | --- | --- | --- | --- |
| Australia | Dynata | 20-Mar | 700 | 46.3 (16.4) | 51.0 | 43.4 | 82.9 | 88.7 |
| China* | Respondi | 09-Apr | 700 | 43.2 (14.3) | 48.9 | 73.1 | 85.8 | 87.4 |
| Germany | Respondi | 23-Mar | 700 | 46.6 (16.0) | 49.9 | 32.7 | 80.8 | 89.2 |
| Spain | Respondi | 22-Mar | 700 | 46.6 (15.0) | 51.1 | 58.1 | 83.6 | 89.8 |
| Spain | Respondi | 06-May | 700 | 46.0 (15.0) | 50.4 | 57.0 | 79.8 | 82.5 |
| France* | BVA | 03-Apr | 3002 | 48.8 (16.5) | 47.5 | 71.1 | 69.7 | 80.7 |
| Italy | Respondi | 22-Mar | 700 | 46.9 (26.1) | 50.4 | 41.3 | 85.3 | 88.2 |
| Japan | Respondi | 10-Apr | 699 | 48.1 (16.4) | 50.9 | 53.3 | 74.5 | 80.1 |
| S. Korea | Respondi | 09-Apr | 700 | 45.3 (15.5) | 49.0 | 70.5 | 85.6 | 88.4 |
| Mexico | Respondi | 21-Mar | 693 | 38.4 (14.2) | 50.5 | 66.4 | 88.1 | 90.3 |
| Mexico | Respondi | 06-May | 700 | 38.7 (14.6) | 51.0 | 75.8 | 73.9 | 75.6 |
| Sweden | Respondi | 28-Mar | 700 | 48.4 (77.3) | 49.1 | 40.3 | 66.3 | 77.2 |
| Sweden | Respondi | 17-Apr | 700 | 45.3 (16.7) | 48.9 | 40.2 | 63.4 | 73.7 |
| UK | Prolific | 19-Mar | 703 | 45.6 (15.7) | 50.9 | 53.9 | 80.4 | 91.7 |
| UK | Prolific | 07-May | 1157 | 45.2 (23.1) | 50.7 | 56.5 | 80.4 | 86.7 |
| UK | Prolific | 06-Jul | 1325 | 44.8 (17.5) | 52.5 | 58.5 | 78.9 | 85.3 |
| UK | Prolific | 18-Sep | 1869 | 38.1 (15.0) | 51.2 | 56.2 | 73.0 | 79.5 |
| UK | Respondi | 07-May | 1150 | 45.6 (16.0) | 52.0 | 43.4 | 78.9 | 84.2 |
| UK* | Respondi | 08-Jun | 500 | 45.9 (15.9) | 53.2 | 39.7 | 79.0 | 83.2 |
| UK | Respondi | 06-Jul | 1326 | 46.0 (24.4) | 51.7 | 44.9 | 80.1 | 84.4 |
| UK | Respondi | 18-Sep | 1855 | 45.7 (19.6) | 51.6 | 42.6 | 75.7 | 79.9 |
| UK | Respondi | 29-Oct | 1744 | 47.1 (23.4) | 52.2 | 42.0 | 72.2 | 76.1 |
| US | Prolific | 19-Mar | 702 | 45.1 (15.9) | 50.6 | 66.8 | 75.7 | 85.7 |
| US | Respondi | 07-May | 700 | 45.7 (26.5) | 51.0 | 59.3 | 74.7 | 80.1 |
| US* | Respondi | 28-Sep | 909 | 44.8 (15.6) | 50.6 | 50.1 | 62.6 | 67.5 |

\*Indicates survey that included vaccine acceptance items but not all model predictor variables (excluded from analyses below).

## Methods

### Participants and procedure

Between March and October 2020, we fielded 25 separate surveys across 12 countries. The majority of samples were recruited through an ISO certified international survey company Respondi (respondi.com). Participants in Australia were recruited through Dynata (dynata.com), and additional US and UK samples were recruited via Prolific (prolific.ac). Quota-based sampling ensured all samples were representative of the country population in terms of age and gender, and, in Prolific samples, ethnicity ^39^. Participants who had previously completed a survey were prevented from completing further surveys, so all our samples represent different individuals. Demographic details for each sample are shown in Table 1. For completeness we include several samples in which vaccine acceptance was measured, but the survey did not always include all the predictor variables used in models presented below. Surveys which did not include all predictor variables are marked with a ‘*’ in Table 1.

All participants were directed via a study link to the Qualtrics platform, and provided informed consent before completing the survey. This study was overseen by the University of Cambridge Psychology Research Ethics Committee (PRE.2020.034).

### Materials

Participants reported their age and gender, level of education (ranging from *No formal education above age 16* to *PhD*), and political orientation (*Very liberal/left wing* to *Very conservative/right wing*). Numeracy was measured as a combined index of the 2-3 item adaptive form of the Berlin Numeracy Test ^40^ and an additional risk literacy item from Lipkus et al.^41^.

Participants completed a widely used measure of general social trust (*Generally speaking, would you say most people can be trusted, or that you can’t be too careful in dealing with people*?)^42^ and a separate measure of prosociality (*To what extent do you think it’s important to do things for the benefit of others and society even if they have some costs to you personally?*). Trust in experts and trust in government were each measured as the combined average of reported trust in three targets (experts: scientists, medical doctors and nurses, and scientific knowledge [Cronbach’s αs .77-.86]; government: politicians, current government, civil servants [αs .73-.90]; all from *Cannot be trusted at all* to *Can be trusted a lot*). We also asked participants to report their trust in several actors with specific regard to the COVID-19 pandemic. Participants reported the extent to which they trust politicians in their country to ‘deal effectively with the pandemic’, and how much they separately trusted the country’s national scientific and medical advisors, independent experts not connected with government, and the WHO to ‘know the best measures to take in the face of the pandemic’ (all from *Not at all* to *Very much*). Personal and government efficacy were captured by items asking participants the extent to which they felt that, respectively, their own actions, and the actions of their country ‘to limit the spread of coronavirus can make a difference’ (*Not at all* to *Very much*). Perceived likelihood of infection was measured as an index of three related items (example: *I will probably get sick with the coronavirus/COVID-19*; αs .71-.89). Participants also reported their level of worry about the virus (from *Not at all worried* to *Very worried*). In a subset of UK samples, we also asked participants about their general attitude towards vaccination, using two items from Lewandowsky et al.’s ^35^ scale (example: *I believe that vaccines are a safe and reliable way to help avert the spread of preventable diseases* [*r*s .83-.87]).

Participants’ vaccine acceptance was measured with the question: ‘*If a vaccine were to be available for the coronavirus/COVID-19 now, would you get vaccinated yourself?*’ (*Yes/No)*. Participants were also asked *‘If a vaccine were to be available for the coronavirus/COVID-19 now: Would you recommend vulnerable friends/family to get vaccinated?*’ (*Yes/No)*. Full item wording for all measures can be found in Table S1.

Surveys were translated from English to other languages by native speakers fluent in English. Multi-item scales (trust in science, trust in government and perceived likelihood of infection) were subjected to multi-group confirmatory factor analysis to establish measurement invariance^43^. All scales exhibited metric invariance based on a criterion of a reduction in CFI no greater than .02 when constraining item factor loadings to be equal across different countries (see Table S2). This more relaxed criterion (compared to the widely used ΔCFI < .01^43^) was applied in light of the recommendations of Rutkowski and Svetina^44^ for analyses with a large number of groups. Metric invariance indicates that effects of the construct in question (but necessarily not latent means) can be compared across groups.

## Results

Figure 1 shows the percentage of participants in each survey who responded that they would be willing to be vaccinated if a COVID-19 vaccine was available, or would recommend a vaccine to vulnerable others, given the options of ‘Yes’ or ‘No’ ^1^. Across all samples, the percentage of respondents who stated they were willing to receive a vaccine ranged from 62.6% (Sweden, April) to 88.1% (Mexico, March), while the percentage of those who said they would recommend a vaccine to vulnerable others ranged from 67.5% (US, September) to 91.7% (UK, March). Descriptively, in every single sample the proportion of respondents stating a willingness to receive a vaccine was lower than the proportion who would recommend it to vulnerable others (*M*_diff_ = −5.79%, SD = 3.00). We also note a trend of decreasing stated acceptance over time: in nearly all countries with multiple samples, vaccine acceptance in any given survey was lower than previous surveys of the same population. For example between March and May, 2020, stated vaccine acceptance among respondents in Mexico dropped from 88.1% to 73.9% (a two-sample proportion test indicated that this difference was statistically significant, 95%CI [-18.4%, −9.9%], z = 6.51, *p* < .001) In the US, stated vaccine acceptance (among participants recruited through online panel provider Respondi) fell more than 12 percentage points, from 74.7% to 62.6%, between May and September, 2020 (95%CI [-16.7%, −7.5%], z = 5.09, *p* < .001)

**Figure 1.**
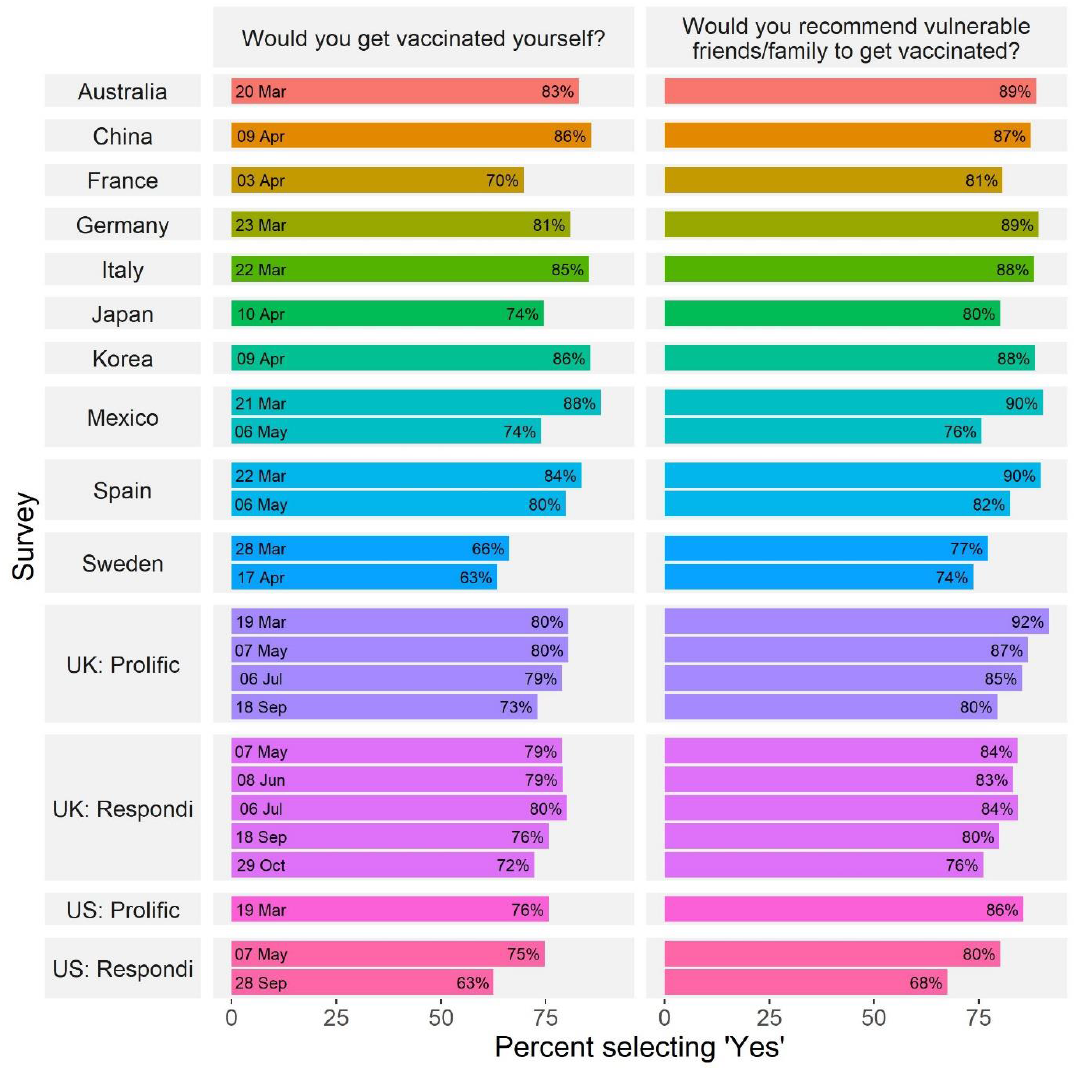
COVID-19 vaccine acceptance across countries and time. Percentage of respondents who stated they were willing to receive or recommend a COVID-19 vaccine across surveys. UK and US samples using different panel providers are reported separately.

We fitted a logistic regression model to data from each sample to identify the correlates of COVID-19 vaccine intentions. Predictors included: demographic variables; an objective measure of numeracy, political ideology; general social trust; prosociality (willingness to ‘do things for the benefit of others and society’ even at personal cost); general trust in medical and scientific experts; general trust in government; specific trust in politicians to manage the pandemic; specific trust in (separately) national science advisors, independent scientists and the WHO to ‘know the best measures to take in the face of the pandemic’; the perceived efficacy of their own and their country’s actions to limit the spread of the virus; perceived likelihood of infection; and, worry about COVID-19 (for details on measures see Methods section and Table S1; descriptive statistics are reported in Tables S3 and S4, ad bivariate correlations in Figure S1). Continuous measures (i.e. all except gender) were scaled and mean centered prior to analysis. Multicollinearity analyses indicated no issues arising from correlated predictors (all variance inflation factor values < 4). To facilitate the interpretation of results we present odds ratios in a heat map format in Figure 2. A full table of model results including confidence intervals can be found in Tables S5 and S6. Results of models predicting vaccine recommendation responses are also presented in supplementary materials (Figure S2, Tables S7 and S8).

**Figure 2.**
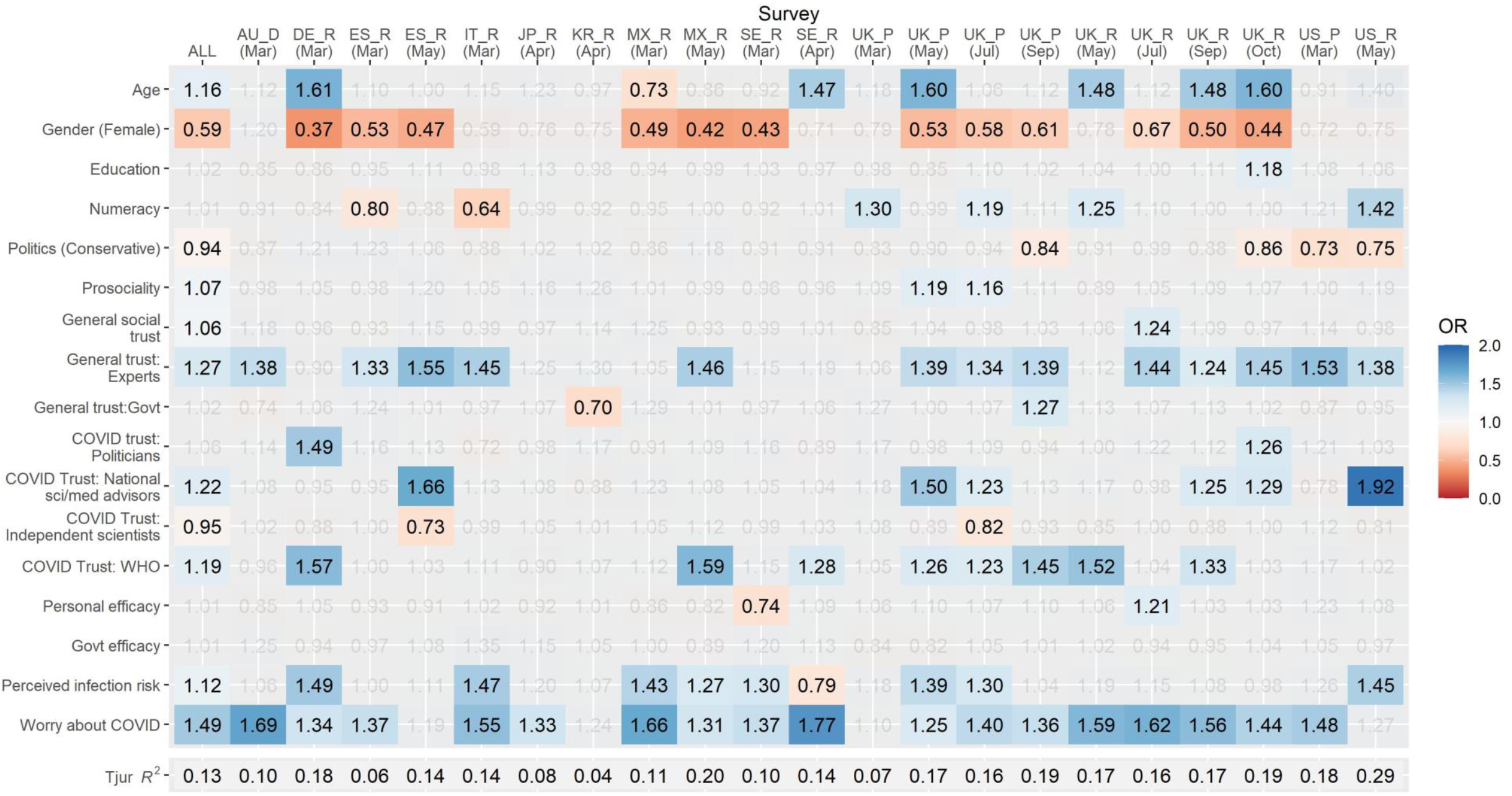
Predictors of vaccine acceptance. Heatmap of odds ratios in logistic regression model predicting stated vaccine acceptance. Columns represent individual samples and rows represent predictors in model. Grey values are non-significant, *p* > .05. Red shading indicates a lower likelihood of reported vaccine acceptance and blue shading a higher likelihood. For space, samples are defined by their two character ISO country code and a letter denoting participant source (D, Dynata; R, Respondi; P, Prolific).

Considering the most consistent predictors of stated vaccine acceptance across samples, we find that in most samples individuals who report a higher level of general trust in experts (OR_pooled_ = 1.27, 95%CI [1.22, 1.33]), or who are more worried about the virus (OR_pooled_ = 1.49, [1.43, 1.55]), are more likely to say that they would accept a vaccine. In Germany, Spain, Mexico, Sweden (March only), and nearly all UK samples, females are generally less likely to say that they would accept a COVID-19 vaccine if available (OR_pooled_ = 0.59, [0.55, 0.64])^2^. We also note that measures of efficacy, both at the personal (OR_pooled_ = 1.01, [0.97, 1.06]) and country level (OR_pooled_ = 1.01, [0.96, 1.07]), were not significantly associated with reported vaccine acceptance in most samples.

Our results reveal a great deal of heterogeneity in the relevance of predictors across countries, but also across time in countries where we conducted multiple surveys. For example, in the United States only a few consistent predictors emerged. Most notably, political conservatism was associated with a lower likelihood to accept a COVID-19 vaccine (OR_USA—Mar_ = 0.73 [0.57, 0.93]; OR_USA—May_ = 0.75, [0.57, 0.99]) whereas trust in experts (OR_USA—Mar_ = 1.53 [1.16, 2.03]; OR_USA—May_ = 1.38, [1.03, 1.84]) and personal worry about the virus (OR_USA—Mar_ = 1.48 [1.17, 1.87]; OR_USA—May_ = 1.27, [0.99 – 1.64]) were associated with increased vaccination intentions. In contrast, in the United Kingdom, additional factors such as the role of age, gender, and prosociality played a significant role. There was also variation over time. For example, although political ideology was not a significant predictor in the UK in May or July, conservatism was associated with lower vaccination intentions from September onwards (ORs 0.84-.88), which may be related to increased polarization. To illustrate the increasing strength of the association between political ideology and vaccine acceptance over time in the UK, in Figure 3 we plot the predicted likelihood of reported vaccine acceptance across the political spectrum (holding all other predictors constant).

**Figure 3.**
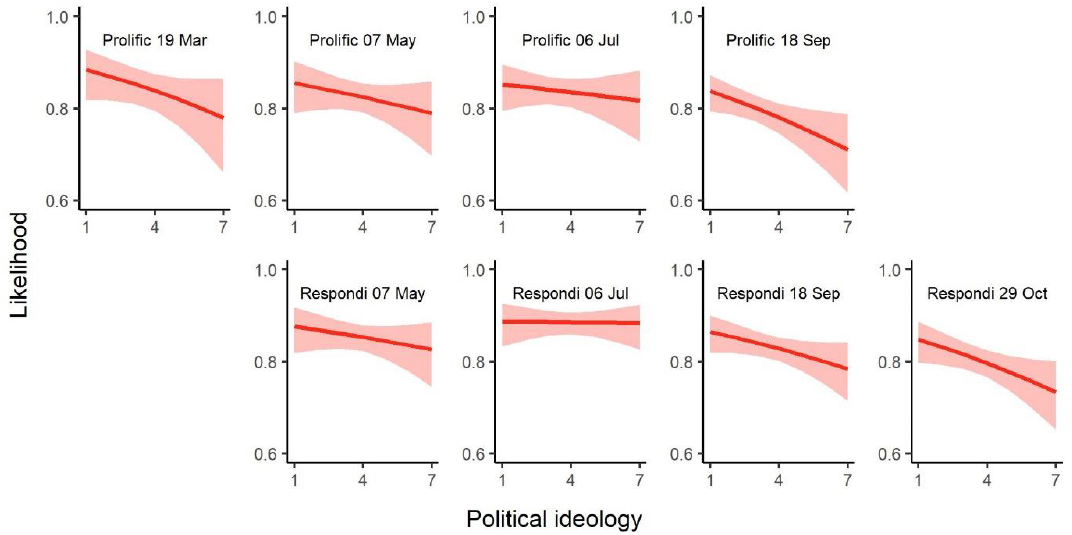
Political ideology and vaccine acceptance in the UK. Predicted likelihood that an individual will accept being vaccinated at varying levels of political ideology (1 = very liberal/left wing, 7 = very conservative/right wing) in UK samples over time.

In the UK, we also report a different pattern of effects when comparing between samples collected via different providers, even where these were collected on the same day (in May, July, and September), were matched on age and gender, and controlling for a range of other demographic variables. This underscores the caution that must be applied when studies generalize results from a single survey sample (particularly an online survey).

In terms of variance explained, the variables in our model explained approximately 10-30% of the variance in the likelihood of vaccine acceptance vs refusal, with the exception of samples recruited in Korea (4%) and Japan (8%).

### Accounting for general vaccine attitudes

To examine the extent to which the effects in our model can be accounted for by a negative perception of vaccines in general, we conducted an additional set of analyses. In our three most recent UK surveys we included a two-item measure of general vaccine attitudes (adapted from Lewandowsky et al.^35^). A comparison of results from models with or without general vaccine attitudes as a predictor is shown as a heat map in Figure 4. Although attitudes toward vaccination increase the explained variance of our model (Δ*R*^2^ 4%-9%) and reveal strong significant effects such that more positive attitudes are associated with increased vaccination intentions (ORs 1.69-2.31; full results in Table S9), the relationships in the original model appear robust and are only minimally attenuated when accounting for generalized attitudes.

**Figure 4.**
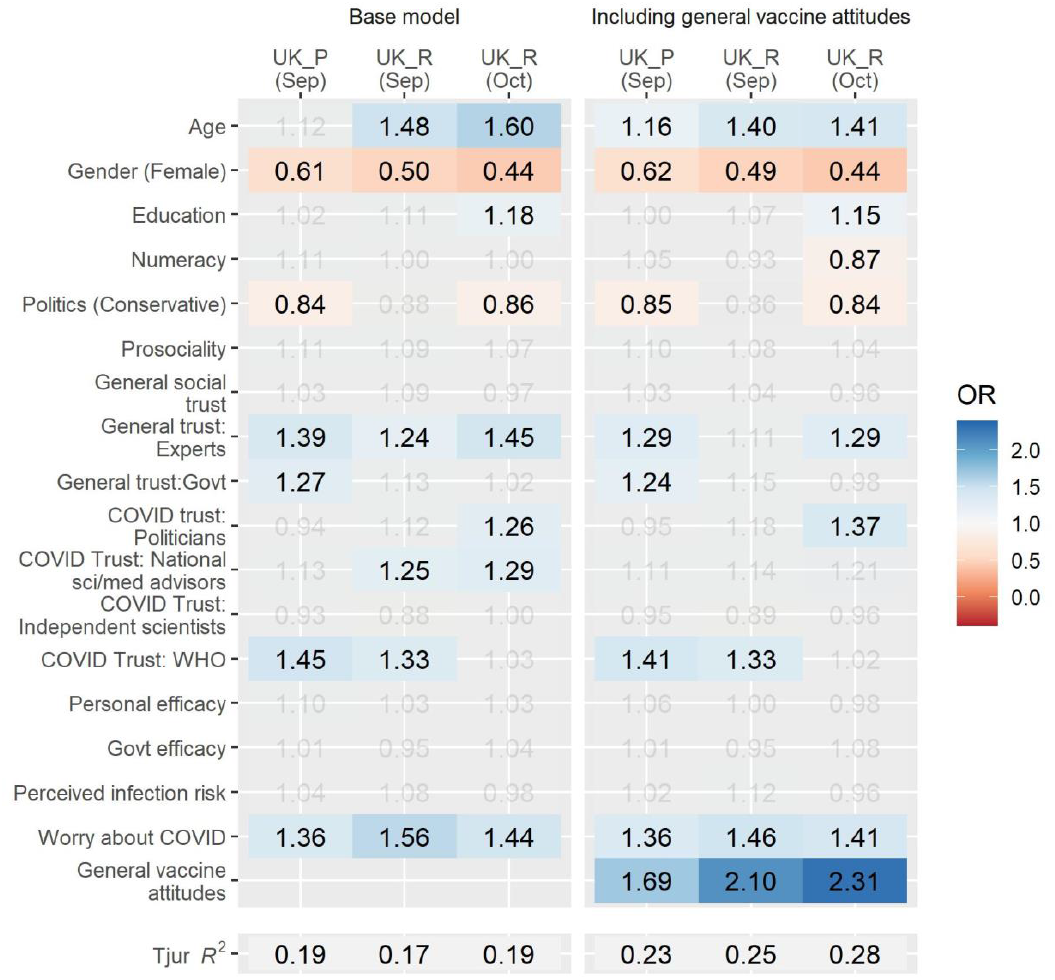
Negative general attitudes towards vaccination do not fully account for relationships in the model. Results of logistic regression models predicting reported COVID-19 vaccine acceptance in UK samples, excluding (left panel) or including (right panel) general vaccine attitudes as a predictor. Odds ratios shown are based on scaled predictors (other than gender). Grey values are non-significant, *p* > .05. For space, samples are defined by a letter denoting participant source (R, Respondi; P, Prolific).

## Discussion

Understanding the psychological determinants of vaccine acceptance and hesitancy is crucial during a global pandemic. Across all countries surveyed, between March and September 2020, a substantial proportion of participants (up to 37% in some countries) said that they would *not* accept a hypothetical COVID-19 vaccine. People were slightly more likely to say that they would recommend it to vulnerable friends and family members. Considering who is more or less likely to report willingness to be vaccinated against COVID-19, being male, expressing general trust in those with scientific or medical expertise, and worrying about the virus are the most consistent correlates of vaccine acceptance across our samples. It is important to note that hesitancy about a COVID-19 vaccine is not purely attributable to people’s attitudes to vaccines in general. Although (in the UK, where we studied it) negative attitudes towards vaccines in general are a significant and important predictor of COVID-19 vaccine refusal, there are clearly additional factors at play in determining public reactions to a COVID-19 vaccine. This broadly aligns with other research indicating that, for many people, there are concerns specifically around the rapid and novel development processes of COVID-19 vaccines and possible safety issues^29,45^. Our multivariate analyses show that the bulk of the burden of trust rests on science and medicine. Accounting for the other factors in our model, we find that trust in government (both generally and regarding COVID-19) and general social trust (i.e. trust in people) are *not* significantly associated with vaccine acceptance in most of our samples.

The fact that we saw only a weak link between stated vaccine acceptance and our measure of prosociality—along with the fact that higher numbers of people said that they’d recommend the vaccine to a vulnerable friend or relative than say they would accept it themselves—suggests that the prosocial nature of vaccines may not be recognized by many people. Recent experimental research has shown that emphasizing the societal benefits of herd immunity (i.e., the need for those who do not see themselves as personally vulnerable to take the vaccine in order to provide protection for those who are) may assist uptake^46^.

The higher reluctance from women to say that they would take a vaccine is in line with other work focusing on acceptance of a potential COVID-19 vaccine^14,15^, and vaccination generally^22^ but has not been adequately explained. Even when general vaccine beliefs are taken into account, however, the gender bias remains. Qualitative work should focus on investigating this further, in order to understand the root of women’s concerns about the COVID-19 vaccine. We see very little effect of our measures of personal or governmental efficacy, but this may be related to the fact that a vaccine against COVID-19 was hypothetical at the time of the surveys and our measures did not directly ask about vaccination.

Another important finding highlighted by our repeated samples is that vaccine acceptance appears to be politicized in the US and is becoming so in the UK. Our US results agree with previous US research focusing on COVID-19 vaccine acceptance^32,47^, which noted that political conservatives are less accepting of potential COVID-19 vaccines. Our UK results align with those of Maher et al, who, through network analysis, show a pattern of attitudinal alignment over time in a small UK sample, resulting in the emergence of a politically conservative faction expressing less trust in scientists, doctors, and vaccines^17^. Although international research has suggested that political conservatism is correlated with anti-vaccination attitudes globally^33^, we did not find that ideology was associated with vaccine acceptance outside of the US and UK. However most other countries were only surveyed in earlier stages of the pandemic (i.e. prior to May, 2020) and we can therefore not say whether they might have followed a similar pattern to the UK as time went on.

It is possible that misinformation susceptibility^48,49^ and conspiracy thinking^50^ underlie the association between ideology and vaccine attitudes to some extent. For example, Motta et al.^51^ find that far right-wing media outlets have disproportionally spread misinformation during the early stages of the pandemic. Susceptibility to misinformation around COVID-19 was also found in prior research to be associated with measures of vaccine hesitancy^49^. There is already a proliferation of conspiracy theories focused on specific COVID-19 vaccines^52,53^. It will be important to tackle these pro-actively through ‘prebunking’ methods to inoculate against misinformation^54,55^.

Finally, we acknowledge that the heterogeneity in our results across time and countries highlights the role that (unmeasured) contextual, country-specific factors play in informing individuals’ vaccination attitudes. As noted by the WHO SAGE working group on vaccine hesitancy, individual factors such as trust and risk perception intersect with contextual influences such as culture, media environments, and information from local leaders^10^. Lastly, our samples were not truly representative of the general population in each country: although they were quota-balanced on gender and age, the population that respond to an online questionnaire will differ from the general population on several significant characteristics. However, the rank ordering of countries on vaccine acceptance in our study is similar to that of Lazarus et al^10^, which were based on a random stratified sampling approach using several online panel providers. This gives us some confidence in the generalizability of our results, and the fact that our samples were generally larger and included more trust-focused questions makes them useful for exploring these important predictors of vaccine attitudes.

In terms of practical considerations, our finding that trust in scientific and medical institutions is one of the strongest predictors of vaccine acceptance highlights the need to work proactively with others from outside of this sphere, such as community and religious leaders^56^ to open a two-way conversation with those who distrust the scientific and medical establishment. Due consideration must also be given to the accessibility^57^, format^58,59^, and transparency^60,61^ of information provided to the public. Future research should continue to evaluate how to most effectively communicate evidence about vaccination, and should seek to more deeply understand the concerns and needs of those who express hesitancy regarding COVID-19 vaccination. As Bhopal^62^, commenting on potential COVID-19 mass vaccination efforts, writes, “Open, honest, factual and sensitively conducted public dialogue is now urgent.”

## Conclusions

Countries around the world face a major evidence communication challenge when it comes to the COVID-19 vaccines that are about to become available. In order to reach a large enough proportion of the population in each country to achieve herd immunity, it is vital to increase in the number of people who are willing to take a vaccine. To achieve this, non-pharmaceutical interventions will need to be deployed^63^, such as communicating trustworthy information about the vaccines via credible sources. In the current research, we have demonstrated across 12 national samples that people’s level of worry about COVID-19 and their trust in experts and medical and scientific institutions are key determinants of potential vaccine acceptance. Future research should confirm these findings in experimental settings. We recommend that empirical studies should continue to be carried out alongside qualitative work with different communities to get a rounded understanding of people’s concerns and misunderstandings. Only by knowing these can we adequately address them and provide people with the information they need to make a decision that will affect not just their own health, but that of their community as well.

## Data Availability

The data and analysis code for this study are available on OSF.

https://osf.io/vgez2/?view_only=8fe81f5fe3f345a99b06edeaba6bd9e1

## Data availability

The data and analysis code for this study are available at: https://osf.io/vgez2/?view_only=8fe81f5fe3f345a99b06edeaba6bd9e1

## Acknowledgments

This study was funded by the Winton Centre for Risk and Evidence Communication which is supported by the David and Claudia Harding Foundation. We would like to thank María del Carmen Climént Palmer, Ban Mutsuhisa, Jin Park, and Giulia Luoni for additional translations, the University of Tokyo for their collaboration, and all the participants and those who helped to administer the study.

## Supplementary material

**Table S1.** Survey items and wording

| Variable | Wording | Response |
| --- | --- | --- |
| Gender | What is your gender? | 0 =Male, 1= Female |
| Age | What is your age? | Age in years |
| Education | Please indicate your highest educational qualification | 1 = No formal education above age 16, 2 = Professional or technical qualifications above age 16, 3 = School education up to age 18, 4 =Degree (Bachelors) or equivalent, 5 = Degree (Masters) or other postgraduate qualification, 6 = Doctorate [In France] 1 = No diploma, 2 =Primary school certificate, 3 = BEPC - Brevet des colleges, 4 = CAP / BEP, 5 = BAC / professional certificate / technical certificate, 6 = BAC +2 and above. |
| Numeracy (summed; range 1-5) | Adaptive Berlin Numeracy test (2-3items, see Cokely et al., 2012 for details).<br>Which represents the highest risk of something happening? | Scores range 1-4<br>1 = '1 in 10' (correct), 2 = '1 in 1000', 3 = '1 in 100' |
| Politics | Where do you feel your political views lie on a spectrum of left wing (or liberal) to right wing (or conservative)? | 1 = Very liberal/left, 7 = Very conservative/right |
| Prosociality | To what extent do you think it's important to do things for the benefit of others and society even if they have some costs to you personally? | 1 = Not at all, 7 = Very much so |
| General social trust | Generally speaking, would you say most people can be trusted, or that you can't be too careful in dealing with people? | 1 = Can't be too careful, 7 = Most people can be trusted |
| General trust: Experts (scale) | How much do you trust each of the following? - Medical doctors and nurses | 1 = Cannot be trusted at all, 5 = Can be trusted a lot |
|  | How much do you trust each of the following? - Scientists | 1 = Cannot be trusted at all, 5 = Can be trusted a lot |
|  | How much do you trust each of the following? - Scientific knowledge | 1 = Cannot be trusted at all , 5 = Can be trusted a lot |
| General trust: Govt (scale) | How much do you trust each of the following? - Civil servants or public officials in the country you are living in | 1 = Cannot be trusted at all , 5 = Can be trusted a lot |
|  | How much do you trust each of the following? - The current government of the country you are living in | 1 = Cannot be trusted at all , 5 = Can be trusted a lot |
|  | How much do you trust each of the following? - Politicians in the country you are living in | 1 = Cannot be trusted at all , 5 = Can be trusted a lot |
| COVID trust: Politicians | How much do you trust the country's politicians to deal effectively with the pandemic? | 1 = Not at all, 7 = Very much |
| COVID Trust: National sci/med advisors | How much do you trust the country's national scientific and medical advisors to know the best measures to take in the face of the pandemic? | 1 = Not at all, 7 = Very much |
| COVID Trust: Independent scientists | How much do you trust experts who are not connected with the government who are commenting on measures planned for the pandemic? | 1 = Not at all, 7 = Very much |
| COVID Trust: WHO | How much do you trust the World Health Organisation to know the best measures to take in the face of the pandemic? | 1 = Not at all, 7 = Very much |
| Personal efficacy | To what extent do you feel that the personal actions you are taking to try to limit the spread of coronavirus make a difference? | 1 = Not at all, 7 = Very much |
| Govt efficacy | To what extent do you feel the actions that your country is taking to limit the spread of coronavirus make a difference? | 1 = Not at all, 7 = Very much |
| Perceived infection risk (scale) | How likely do you think it is that you will be directly and personally affected by the following in the next 6 months? - Catching the coronavirus/COVID-19 | 1= Not at all likely, 7 = Very likely |
|  | How likely do you think it is that your friends and family in the country you are currently living in will be directly affected by the following in the next 6 months? - Catching the coronavirus/COVID-19 | 1 = Not at all likely, 7 = Very likely |
|  | How much do you agree or disagree with the following statements? - I will probably get sick with the coronavirus/COVID-19. | 1 = Strongly disagree, 5 = Strongly agree (rescaled) |
| Worry about COVID | How worried are you personally about the following issues at present? - Coronavirus/COVID-19 | 1 = Not at all worried, 7 = Very worried |
| Vaccine - acceptance | If a vaccine were to be available for the coronavirus/COVID-19 now: - Would you get vaccinated yourself? | 0 = No, 1 = Yes |
| Vaccine – recommend to others | If a vaccine were to be available for the coronavirus/COVID-19 now: - Would you recommend vulnerable friends/family to get vaccinated? | 0 = No, 1 = Yes |
| General vaccine attitudes | Please let us know how much you agree or disagree with the following statements about vaccines in general: - I believe that vaccines are a safe and reliable way to help avert the spread of preventable diseases | 1 = Strongly disagree, 5 = Strongly agree |
|  | Please let us know how much you agree or disagree with the following statements about vaccines in general: - Vaccinations are one of the most significant contributions to public health | 1 = Strongly disagree, 5 = Strongly agree |

**Table S2.** Results of measurement invariance analyses

Results of measurement invariance analyses
| Model | Constraints | *Df* | $\chi^2$ | $\Delta\chi^2$ | $\Delta Df$ | CFI | RMSEA | SRMR | $\Delta CFI$ | $\Delta RMSEA$ | $\Delta SRMR$ |
| --- | --- | --- | --- | --- | --- | --- | --- | --- | --- | --- | --- |
| Trust in experts | Configural model | 0 | 0 | - | - | 1 | 0 | 0 | - | - | - |
|  | Loadings | 18 | 74.49 | 74.49*** | 18 | 0.998 | 0.039 | 0.017 | 0.002 | 0.039 | 0.017 |
|  | Intercepts | 36 | 488.11 | 413.62*** | 18 | 0.981 | 0.079 | 0.035 | 0.017 | 0.039 | 0.018 |
|  | Means | 45 | 983.43 | 495.32*** | 9 | 0.960 | 0.102 | 0.067 | 0.021 | 0.023 | 0.032 |
| Trust in government | Configural model | 0 | 0 | - | - | 1 | 0 | 0 | - | - | - |
|  | Loadings | 18 | 447.63 | 447.63*** | 18 | 0.981 | 0.109 | 0.049 | 0.019 | 0.109 | 0.049 |
|  | Intercepts | 36 | 1628.51 | 1180.88*** | 18 | 0.931 | 0.148 | 0.070 | 0.051 | 0.039 | 0.020 |
|  | Means | 45 | 2318.57 | 690.06*** | 9 | 0.901 | 0.158 | 0.104 | 0.030 | 0.010 | 0.035 |
| Perceived likelihood of infection | Configural model | 0 | 0 | - | - | 1 | 0 | 0 | - | - | - |
|  | Loadings | 18 | 124.79 | 124.79*** | 18 | 0.996 | 0.054 | 0.018 | 0.004 | 0.054 | 0.018 |
|  | Intercepts | 36 | 826.98 | 702.19*** | 18 | 0.968 | 0.104 | 0.047 | 0.028 | 0.050 | 0.028 |
|  | Means | 45 | 1036.54 | 209.56*** | 9 | 0.960 | 0.105 | 0.061 | 0.008 | 0.000 | 0.015 |

\*\*\* $p < .001$ , chi-square difference test

**Table S3.** Descriptive statistics for all samples (Mean (SD)), excluding US and UK samples (see Table S4)

|  | ALL | AU_D<br>(Mar) | CN_R<br>(Apr) | DE_R<br>(Mar) | ES_R<br>(Mar) | ES_R<br>(May) | FR_B<br>(Apr) | IT_R<br>(Mar) | JP_R<br>(Apr) | KR_R<br>(Apr) | MX_R<br>(Mar) | MX_R<br>(May) | SE_R<br>(Mar) | SE_R<br>(Apr) |
| --- | --- | --- | --- | --- | --- | --- | --- | --- | --- | --- | --- | --- | --- | --- |
| Age | 45.27<br>(22.29) | 46.30<br>(16.44) | 43.21<br>(14.26) | 46.61<br>(16.00) | 46.64<br>(15.03) | 46.00<br>(15.03) | 48.79<br>(16.53) | 46.95<br>(26.06) | 48.08<br>(16.35) | 45.34<br>(15.51) | 38.39<br>(14.24) | 38.68<br>(14.56) | 48.41<br>(77.28) | 45.31<br>(16.74) |
| Gender (Female) | 0.51<br>( 0.50) | 0.51<br>( 0.50) | 0.49<br>( 0.50) | 0.50<br>( 0.50) | 0.51<br>( 0.50) | 0.50<br>( 0.50) | 0.48<br>( 0.50) | 0.50<br>( 0.50) | 0.51<br>( 0.50) | 0.49<br>( 0.50) | 0.50<br>( 0.50) | 0.51<br>( 0.50) | 0.49<br>( 0.50) | 0.49<br>( 0.50) |
| Education | 3.42<br>( 1.13) | 3.17<br>( 1.12) | 3.64<br>( 0.88) | 3.07<br>( 1.19) | 3.59<br>( 1.08) | 3.60<br>( 1.07) | 5.00<sup>a</sup><br>(1.12) | 3.51<br>( 1.23) | 3.56<br>( 0.81) | 3.76<br>( 0.79) | 3.55<br>( 0.94) | 3.74<br>( 0.88) | 3.30<br>( 1.06) | 3.34<br>( 1.01) |
| Numeracy | 2.66<br>( 1.11) | 2.41<br>( 1.06) | 2.83<br>( 1.25) | 2.53<br>( 1.12) | 2.39<br>( 1.03) | 2.42<br>( 0.97) | 2.18<br>( 0.70) | 2.13<br>( 0.71) | 2.77<br>( 1.25) | 2.60<br>( 1.06) | 2.16<br>( 0.90) | 2.31<br>( 0.95) | 2.52<br>( 1.23) | 2.57<br>( 1.21) |
| Politics (Conservative) | 3.74<br>( 1.41) | 3.83<br>( 1.30) | - | 3.79<br>( 1.19) | 3.50<br>( 1.43) | 3.44<br>( 1.50) | - | 3.87<br>( 1.45) | 4.09<br>( 1.09) | 3.88<br>( 1.20) | 3.65<br>( 1.31) | 3.57<br>( 1.33) | 3.92<br>( 1.59) | 3.88<br>( 1.61) |
| Prosociality | 5.21<br>( 1.36) | 5.23<br>( 1.36) | 5.54<br>( 1.19) | 4.97<br>( 1.42) | 5.74<br>( 1.21) | 5.35<br>( 1.34) | 5.23<br>( 1.41) | 5.76<br>( 1.38) | 4.74<br>( 1.42) | 4.40<br>( 1.31) | 5.34<br>( 1.61) | 5.26<br>( 1.53) | 4.87<br>( 1.43) | 4.63<br>( 1.48) |
| General social trust | 3.66<br>( 1.71) | 3.95<br>( 1.67) | 4.96<br>( 1.67) | 3.61<br>( 1.65) | 3.47<br>( 1.87) | 3.29<br>( 1.77) | 2.98<br>( 1.62) | 3.70<br>( 1.61) | 3.85<br>( 1.48) | 3.97<br>( 1.50) | 2.81<br>( 1.84) | 2.94<br>( 1.84) | 3.73<br>( 1.73) | 3.85<br>( 1.73) |
| General trust: Experts | 3.97<br>( 0.77) | 3.97<br>( 0.79) | 4.26<br>( 0.61) | 3.90<br>( 0.74) | 4.19<br>( 0.72) | 4.09<br>( 0.76) | 3.76<br>( 0.81) | 4.02<br>( 0.74) | 3.51<br>( 0.72) | 3.74<br>( 0.68) | 4.05<br>( 0.85) | 4.10<br>( 0.83) | 3.90<br>( 0.75) | 3.85<br>( 0.75) |
| General trust: Govt | 2.64<br>( 0.91) | 2.96<br>( 0.93) | - | 3.14<br>( 0.92) | 2.75<br>( 0.94) | 2.54<br>( 0.89) | 2.48<br>( 0.90) | 2.89<br>( 0.91) | 2.46<br>( 0.84) | 2.65<br>( 0.81) | 2.28<br>( 1.01) | 2.35<br>( 1.02) | 3.04<br>( 1.00) | 3.00<br>( 1.01) |
| COVID trust: Politicians | 3.48<br>( 1.87) | 4.44<br>( 1.75) | - | 4.80<br>( 1.65) | 4.01<br>( 1.88) | 3.45<br>( 1.91) | 3.33<br>( 1.78) | 4.35<br>( 1.74) | 3.03<br>( 1.60) | 4.24<br>( 1.63) | 3.22<br>( 1.99) | 3.51<br>( 1.96) | 4.13<br>( 1.78) | 4.24<br>( 1.83) |
| COVID Trust: National sci/med advisors | 4.91<br>( 1.60) | 5.34<br>( 1.41) | - | 5.40<br>( 1.43) | 5.45<br>( 1.39) | 4.92<br>( 1.65) | 4.46<br>( 1.73) | 5.42<br>( 1.40) | 3.90<br>( 1.51) | 5.15<br>( 1.37) | 5.18<br>( 1.69) | 5.28<br>( 1.60) | 4.78<br>( 1.67) | 4.93<br>( 1.66) |
| COVID Trust: Independent scientists | 4.66<br>( 1.55) | 4.77<br>( 1.49) | - | 5.11<br>( 1.38) | 5.15<br>( 1.39) | 4.83<br>( 1.55) | 4.62<br>( 1.52) | 4.80<br>( 1.45) | 3.73<br>( 1.51) | 5.03<br>( 1.26) | 5.02<br>( 1.68) | 4.91<br>( 1.65) | 4.66<br>( 1.52) | 4.55<br>( 1.48) |
| COVID Trust: WHO | 4.84<br>( 1.68) | 5.19<br>( 1.55) | - | 5.16<br>( 1.52) | 5.46<br>( 1.42) | 4.88<br>( 1.62) | 4.80<br>( 1.61) | 5.25<br>( 1.48) | 3.12<br>( 1.55) | 3.96<br>( 1.51) | 5.80<br>( 1.45) | 5.58<br>( 1.62) | 5.05<br>( 1.45) | 4.81<br>( 1.55) |
| Personal efficacy | 5.22<br>( 1.48) | 5.14<br>( 1.45) | - | 5.24<br>( 1.41) | 5.31<br>( 1.47) | 5.14<br>( 1.47) | 5.20<br>( 1.47) | 5.31<br>( 1.46) | 4.26<br>( 1.42) | 5.42<br>( 1.24) | 5.36<br>( 1.66) | 5.56<br>( 1.53) | 5.20<br>( 1.50) | 5.26<br>( 1.45) |
| Govt efficacy | 3.86<br>( 1.78) | 4.48<br>( 1.63) | - | 4.68<br>( 1.47) | 4.21<br>( 1.77) | 4.11<br>( 1.83) | 4.22<br>( 1.68) | 4.60<br>( 1.60) | 3.21<br>( 1.52) | 5.08<br>( 1.52) | 3.82<br>( 2.06) | 4.39<br>( 1.86) | 4.30<br>( 1.73) | 4.41<br>( 1.67) |
| Perceived infection risk | 4.17<br>( 1.32) | 4.16<br>( 1.40) | 3.26<br>( 1.37) | 4.13<br>( 1.34) | 4.46<br>( 1.23) | 4.38<br>( 1.26) | 4.19<br>( 1.24) | 3.93<br>( 1.30) | 4.48<br>( 1.20) | 4.37<br>( 1.22) | 4.16<br>( 1.47) | 4.29<br>( 1.43) | 4.30<br>( 1.37) | 4.37<br>( 1.29) |
| Worry about COVID | 5.56<br>( 1.52) | 5.56<br>( 1.51) | 5.37<br>( 1.57) | 5.66<br>( 1.49) | 6.25<br>( 1.17) | 6.11<br>( 1.26) | 5.63<br>( 1.43) | 6.08<br>( 1.27) | 5.83<br>( 1.28) | 5.59<br>( 1.29) | 5.92<br>( 1.46) | 6.06<br>( 1.35) | 5.27<br>( 1.55) | 4.98<br>( 1.66) |
| Vaccine – acceptance | 0.76<br>( 0.43) | 0.83<br>( 0.38) | 0.86<br>( 0.35) | 0.81<br>( 0.39) | 0.84<br>( 0.37) | 0.80<br>( 0.40) | 0.70<br>( 0.46) | 0.85<br>( 0.35) | 0.74<br>( 0.44) | 0.86<br>( 0.35) | 0.88<br>( 0.32) | 0.74<br>( 0.44) | 0.66<br>( 0.47) | 0.63<br>( 0.48) |
| Vaccine – recommend to vulnerable others | 0.82<br>( 0.38) | 0.89<br>( 0.32) | 0.87<br>( 0.33) | 0.89<br>( 0.31) | 0.90<br>( 0.30) | 0.82<br>( 0.38) | 0.81<br>( 0.40) | 0.88<br>( 0.32) | 0.80<br>( 0.40) | 0.88<br>( 0.32) | 0.90<br>( 0.30) | 0.76<br>( 0.43) | 0.77<br>( 0.42) | 0.74<br>( 0.44) |
| General vaccine attitudes | 4.05<br>( 1.11) | - | - | - | - | - | - | - | - | - | - | - | - | - |

<sup>a</sup> Education item in France differed from other surveys – see Table S1.

**Table S4.** Descriptive statistics for all US and UK samples (Mean (SD))

|  | UK_P<br>(Mar) | UK_P<br>(May) | UK_P<br>(Jul) | UK_P<br>(Sep) | UK_R<br>(May) | UK_R<br>(Jun) | UK_R<br>(Jul) | UK_R<br>(Sep) | UK_R<br>(Oct) | US_P<br>(Mar) | US_R<br>(May) | US_R<br>(Sep) |
| --- | --- | --- | --- | --- | --- | --- | --- | --- | --- | --- | --- | --- |
| Age | 45.63<br>(15.69) | 45.22<br>(23.08) | 44.76<br>(17.55) | 38.14<br>(15.01) | 45.64<br>(15.99) | 45.90<br>(15.87) | 46.01<br>(24.36) | 45.75<br>(19.58) | 47.13<br>(23.44) | 45.09<br>(15.90) | 45.73<br>(26.53) | 44.76<br>(15.60) |
| Gender (Female) | 0.51<br>( 0.50) | 0.51<br>( 0.50) | 0.53<br>( 0.50) | 0.52<br>( 0.50) | 0.52<br>( 0.50) | 0.53<br>( 0.50) | 0.52<br>( 0.50) | 0.52<br>( 0.50) | 0.52<br>( 0.50) | 0.51<br>( 0.50) | 0.51<br>( 0.50) | 0.51<br>( 0.50) |
| Education | 3.45<br>( 1.17) | 3.50<br>( 1.14) | 3.58<br>( 1.14) | 3.54<br>( 1.10) | 3.17<br>( 1.27) | - | 3.20<br>( 1.23) | 3.15<br>( 1.28) | 3.10<br>( 1.23) | 3.87<br>( 0.88) | 3.70<br>( 0.90) | - |
| Numeracy | 3.22<br>( 1.17) | 3.23<br>( 1.14) | 3.04<br>( 1.06) | 3.24<br>( 1.15) | 2.64<br>( 1.14) | 2.74<br>( 1.10) | 2.61<br>( 1.07) | 2.60<br>( 1.07) | 2.78<br>( 1.11) | 3.14<br>( 1.13) | 2.76<br>( 1.14) | 2.58<br>( 1.12) |
| Politics (Conservative) | 3.69<br>( 1.43) | 3.67<br>( 1.39) | 3.58<br>( 1.36) | 3.37<br>( 1.36) | 3.90<br>( 1.35) | 3.89<br>( 1.19) | 3.90<br>( 1.33) | 3.84<br>( 1.36) | 3.83<br>( 1.32) | 3.22<br>( 1.65) | 3.92<br>( 1.69) | 4.07<br>( 1.65) |
| Prosociality | 5.50<br>( 1.07) | 5.36<br>( 1.19) | 5.32<br>( 1.16) | 5.42<br>( 1.17) | 5.12<br>( 1.33) | 5.25<br>( 1.29) | 5.03<br>( 1.29) | 5.08<br>( 1.39) | 5.38<br>( 1.36) | 5.43<br>( 1.28) | 5.05<br>( 1.36) | 5.02<br>( 1.44) |
| General social trust | 4.04<br>( 1.59) | 4.12<br>( 1.55) | 4.11<br>( 1.55) | 3.69<br>( 1.56) | 3.74<br>( 1.71) | 3.58<br>( 1.70) | 3.86<br>( 1.59) | 3.68<br>( 1.68) | 3.66<br>( 1.64) | 4.01<br>( 1.68) | 3.79<br>( 1.73) | 3.47<br>( 1.84) |
| General trust: Experts | 4.24<br>( 0.66) | 4.11<br>( 0.63) | 4.14<br>( 0.66) | 4.17<br>( 0.68) | 3.89<br>( 0.79) | 3.88<br>( 0.81) | 3.92<br>( 0.76) | 3.90<br>( 0.78) | 3.92<br>( 0.77) | 4.22<br>( 0.73) | 3.96<br>( 0.77) | 3.89<br>( 0.83) |
| General trust: Govt | 2.82<br>( 0.85) | 2.80<br>( 0.82) | 2.60<br>( 0.82) | 2.44<br>( 0.81) | 2.82<br>( 0.87) | 2.64<br>( 0.87) | 2.70<br>( 0.88) | 2.60<br>( 0.90) | 2.55<br>( 0.86) | 2.55<br>( 0.79) | 2.68<br>( 0.83) | 2.52<br>( 0.88) |
| COVID trust: Politicians | 3.81<br>( 1.78) | 3.80<br>( 1.81) | 3.16<br>( 1.80) | 2.57<br>( 1.65) | 4.00<br>( 1.86) | 3.38<br>( 1.70) | 3.60<br>( 1.83) | 3.23<br>( 1.86) | 3.04<br>( 1.79) | 3.06<br>( 1.74) | 3.11<br>( 1.77) | 2.93<br>( 1.81) |
| COVID Trust: National sci/med advisors | 5.27<br>( 1.47) | 5.13<br>( 1.41) | 5.12<br>( 1.47) | 4.88<br>( 1.58) | 4.94<br>( 1.57) | 4.58<br>( 1.51) | 4.88<br>( 1.53) | 4.66<br>( 1.61) | 4.60<br>( 1.66) | 5.46<br>( 1.41) | 5.15<br>( 1.55) | - |
| COVID Trust: Independent scientists | 4.88<br>( 1.48) | 4.59<br>( 1.44) | 4.74<br>( 1.48) | 4.77<br>( 1.60) | 4.46<br>( 1.52) | - | 4.54<br>( 1.52) | 4.40<br>( 1.61) | 4.28<br>( 1.63) | 5.16<br>( 1.48) | 4.72<br>( 1.60) | - |
| COVID Trust: WHO | 5.59<br>( 1.40) | 4.97<br>( 1.55) | 5.02<br>( 1.62) | 4.77<br>( 1.70) | 4.76<br>( 1.69) | 4.46<br>( 1.69) | 4.72<br>( 1.66) | 4.50<br>( 1.69) | 4.44<br>( 1.71) | 5.62<br>( 1.55) | 4.57<br>( 1.90) | - |
| Personal efficacy | 5.04<br>( 1.39) | 5.59<br>( 1.26) | 5.47<br>( 1.35) | 5.12<br>( 1.48) | 5.36<br>( 1.48) | 5.13<br>( 1.45) | 5.30<br>( 1.45) | 5.09<br>( 1.52) | 5.03<br>( 1.52) | 5.25<br>( 1.45) | 5.32<br>( 1.47) | 5.14<br>( 1.57) |
| Govt efficacy | 3.86<br>( 1.75) | 3.85<br>( 1.70) | 3.48<br>( 1.72) | 3.03<br>( 1.62) | 4.13<br>( 1.74) | 3.66<br>( 1.61) | 3.88<br>( 1.75) | 3.58<br>( 1.73) | 3.36<br>( 1.70) | 3.28<br>( 1.80) | 3.76<br>( 1.76) | 3.25<br>( 1.86) |
| Perceived infection risk | 4.89<br>( 1.32) | 4.26<br>( 1.24) | 3.96<br>( 1.24) | 4.26<br>( 1.30) | 4.14<br>( 1.22) | 3.94<br>( 1.25) | 3.86<br>( 1.25) | 4.13<br>( 1.28) | 4.27<br>( 1.24) | 3.98<br>( 1.52) | 3.91<br>( 1.38) | 4.11<br>( 1.38) |
| Worry about COVID | 5.80<br>( 1.36) | 5.72<br>( 1.40) | 5.28<br>( 1.52) | 5.36<br>( 1.58) | 5.60<br>( 1.51) | 5.34<br>( 1.57) | 5.30<br>( 1.60) | 5.39<br>( 1.61) | 5.39<br>( 1.63) | 5.49<br>( 1.58) | 5.58<br>( 1.60) | 5.43<br>( 1.72) |
| Vaccine – acceptance | 0.80<br>( 0.40) | 0.80<br>( 0.40) | 0.79<br>( 0.41) | 0.73<br>( 0.44) | 0.79<br>( 0.41) | 0.79<br>( 0.41) | 0.80<br>( 0.40) | 0.76<br>( 0.43) | 0.72<br>( 0.45) | 0.76<br>( 0.43) | 0.75<br>( 0.44) | 0.63<br>( 0.48) |
| Vaccine – recommend to vulnerable others | 0.92<br>( 0.28) | 0.87<br>( 0.34) | 0.85<br>( 0.36) | 0.80<br>( 0.40) | 0.84<br>( 0.36) | 0.83<br>( 0.38) | 0.84<br>( 0.36) | 0.80<br>( 0.40) | 0.76<br>( 0.43) | 0.86<br>( 0.35) | 0.80<br>( 0.40) | 0.68<br>( 0.47) |
| General vaccine attitudes | - | - | - | 4.21<br>( 1.10) | - | - | - | 3.90<br>( 1.14) | 4.05<br>( 1.06) | - | - | - |

**Table S5.** Full logistic regression results from model predicting vaccine acceptance, excluding UK and US samples (shown in Table S6)

|  | ALL<br>*OR* | ALL (-UK)<br>*OR* | AU_D (Mar)<br>*OR* | DE_R (Mar)<br>*OR* | ES_R (Mar)<br>*OR* | ES_R (May)<br>*OR* | IT_R (Mar)<br>*OR* | JP_R (Apr)<br>*OR* | KR_R (Apr)<br>*OR* | MX_R (Mar)<br>*OR* | MX_R (May)<br>*OR* | SE_R (Mar)<br>*OR* | SE_R (Apr)<br>*OR* |
| --- | --- | --- | --- | --- | --- | --- | --- | --- | --- | --- | --- | --- | --- |
| (Intercept) | 5.23 ***<br>[4.94 – 5.54] | 5.22 ***<br>[4.79 – 5.69] | 5.59 ***<br>[4.08 – 7.83] | 9.38 ***<br>[6.51 – 13.96] | 8.10 ***<br>[5.74 – 11.79] | 7.35 ***<br>[5.28 – 10.50] | 11.73 ***<br>[7.54 – 19.18] | 3.70 ***<br>[2.78 – 4.99] | 7.89 ***<br>[5.66 – 11.31] | 15.24 ***<br>[9.74 – 25.20] | 5.43 ***<br>[3.99 – 7.55] | 3.17 ***<br>[2.45 – 4.14] | 2.20 ***<br>[1.71 – 2.86] |
| Age | 1.16 ***<br>[1.10 – 1.23] | 1.00<br>[0.95 – 1.07] | 1.12<br>[0.88 – 1.42] | 1.61 ***<br>[1.27 – 2.06] | 1.10<br>[0.87 – 1.38] | 1.00<br>[0.80 – 1.25] | 1.15<br>[0.79 – 1.91] | 1.23<br>[1.00 – 1.52] | 0.97<br>[0.77 – 1.23] | 0.73 *<br>[0.55 – 0.96] | 0.86<br>[0.70 – 1.05] | 0.92<br>[0.56 – 1.09] | 1.47 ***<br>[1.21 – 1.79] |
| Gender (Female)<sup>a</sup> | 0.59 ***<br>[0.55 – 0.64] | 0.61 ***<br>[0.55 – 0.69] | 1.20<br>[0.76 – 1.88] | 0.37 ***<br>[0.23 – 0.59] | 0.53 **<br>[0.33 – 0.83] | 0.47 ***<br>[0.30 – 0.72] | 0.59<br>[0.34 – 1.03] | 0.76<br>[0.50 – 1.13] | 0.75<br>[0.48 – 1.19] | 0.49 *<br>[0.27 – 0.87] | 0.42 ***<br>[0.28 – 0.62] | 0.43 ***<br>[0.30 – 0.62] | 0.71<br>[0.49 – 1.03] |
| Education | 1.02<br>[0.98 – 1.06] | 1.00<br>[0.94 – 1.05] | 0.85<br>[0.67 – 1.07] | 0.86<br>[0.68 – 1.08] | 0.95<br>[0.77 – 1.18] | 1.11<br>[0.90 – 1.36] | 0.98<br>[0.74 – 1.30] | 1.13<br>[0.93 – 1.39] | 0.98<br>[0.78 – 1.23] | 0.94<br>[0.70 – 1.25] | 0.99<br>[0.81 – 1.20] | 1.03<br>[0.86 – 1.23] | 0.97<br>[0.81 – 1.16] |
| Numeracy | 1.01<br>[0.97 – 1.05] | 0.93 *<br>[0.88 – 0.99] | 0.91<br>[0.73 – 1.15] | 0.84<br>[0.67 – 1.05] | 0.80 *<br>[0.65 – 0.99] | 0.88<br>[0.72 – 1.09] | 0.64 **<br>[0.48 – 0.84] | 0.99<br>[0.81 – 1.23] | 0.92<br>[0.73 – 1.16] | 0.95<br>[0.73 – 1.25] | 1.00<br>[0.82 – 1.22] | 0.92<br>[0.77 – 1.11] | 1.01<br>[0.84 – 1.22] |
| Politics (Conservative) | 0.94 **<br>[0.90 – 0.98] | 0.95<br>[0.90 – 1.00] | 0.87<br>[0.68 – 1.11] | 1.21<br>[0.96 – 1.52] | 1.23<br>[0.98 – 1.55] | 1.06<br>[0.84 – 1.33] | 0.88<br>[0.66 – 1.18] | 1.02<br>[0.84 – 1.25] | 1.02<br>[0.80 – 1.30] | 0.86<br>[0.65 – 1.14] | 1.18<br>[0.97 – 1.44] | 0.91<br>[0.76 – 1.10] | 0.91<br>[0.75 – 1.10] |
| Prosociality | 1.07 **<br>[1.03 – 1.11] | 1.06 *<br>[1.00 – 1.13] | 0.98<br>[0.75 – 1.26] | 1.05<br>[0.82 – 1.35] | 0.98<br>[0.78 – 1.24] | 1.20<br>[0.96 – 1.50] | 1.05<br>[0.78 – 1.39] | 1.16<br>[0.93 – 1.45] | 1.26<br>[0.98 – 1.62] | 1.01<br>[0.76 – 1.32] | 0.99<br>[0.81 – 1.22] | 0.96<br>[0.79 – 1.16] | 0.96<br>[0.79 – 1.16] |
| General social trust | 1.06 **<br>[1.02 – 1.11] | 1.06<br>[1.00 – 1.13] | 1.18<br>[0.91 – 1.52] | 0.96<br>[0.75 – 1.24] | 0.93<br>[0.73 – 1.17] | 1.15<br>[0.92 – 1.44] | 0.99<br>[0.74 – 1.33] | 0.97<br>[0.77 – 1.22] | 1.14<br>[0.87 – 1.48] | 1.25<br>[0.94 – 1.70] | 0.93<br>[0.76 – 1.14] | 0.99<br>[0.81 – 1.20] | 1.01<br>[0.83 – 1.23] |
| General trust: Experts | 1.27 ***<br>[1.22 – 1.33] | 1.26 ***<br>[1.18 – 1.35] | 1.38 *<br>[1.06 – 1.80] | 0.90<br>[0.66 – 1.21] | 1.33 *<br>[1.02 – 1.73] | 1.55 ***<br>[1.23 – 1.97] | 1.45 *<br>[1.05 – 2.01] | 1.25<br>[0.97 – 1.60] | 1.30<br>[1.00 – 1.71] | 1.05<br>[0.77 – 1.40] | 1.46 **<br>[1.16 – 1.84] | 1.15<br>[0.91 – 1.44] | 1.19<br>[0.95 – 1.49] |
| General trust: Govt | 1.02<br>[0.96 – 1.07] | 0.93<br>[0.86 – 1.01] | 0.74<br>[0.54 – 1.00] | 1.06<br>[0.77 – 1.47] | 1.24<br>[0.91 – 1.70] | 1.01<br>[0.76 – 1.34] | 0.97<br>[0.67 – 1.39] | 1.07<br>[0.81 – 1.40] | 0.70 *<br>[0.52 – 0.93] | 1.29<br>[0.90 – 1.89] | 1.01<br>[0.78 – 1.31] | 0.97<br>[0.75 – 1.26] | 1.06<br>[0.79 – 1.42] |
| COVID trust: Politicians | 1.06<br>[1.00 – 1.13] | 1.04<br>[0.94 – 1.13] | 1.14<br>[0.78 – 1.66] | 1.49 *<br>[1.02 – 2.20] | 1.16<br>[0.83 – 1.63] | 1.13<br>[0.81 – 1.57] | 0.72<br>[0.45 – 1.12] | 0.98<br>[0.67 – 1.41] | 1.17<br>[0.86 – 1.58] | 0.91<br>[0.61 – 1.36] | 1.09<br>[0.80 – 1.47] | 1.16<br>[0.85 – 1.59] | 0.89<br>[0.64 – 1.23] |
| COVID Trust: National<br>sci/med advisors | 1.22 ***<br>[1.15 – 1.28] | 1.22 ***<br>[1.12 – 1.32] | 1.08<br>[0.76 – 1.52] | 0.95<br>[0.64 – 1.42] | 0.95<br>[0.69 – 1.31] | 1.66 ***<br>[1.24 – 2.24] | 1.13<br>[0.72 – 1.78] | 1.08<br>[0.79 – 1.48] | 0.88<br>[0.63 – 1.24] | 1.23<br>[0.89 – 1.70] | 1.08<br>[0.82 – 1.42] | 1.05<br>[0.79 – 1.41] | 1.04<br>[0.77 – 1.41] |
| COVID Trust:<br>Independent scientists | 0.95 *<br>[0.90 – 0.99] | 1.00<br>[0.93 – 1.07] | 1.02<br>[0.76 – 1.34] | 0.88<br>[0.64 – 1.21] | 1.00<br>[0.76 – 1.31] | 0.73 *<br>[0.56 – 0.95] | 0.99<br>[0.70 – 1.37] | 1.05<br>[0.78 – 1.41] | 1.01<br>[0.74 – 1.37] | 1.05<br>[0.78 – 1.42] | 1.12<br>[0.89 – 1.41] | 0.99<br>[0.78 – 1.24] | 1.03<br>[0.83 – 1.27] |
| COVID Trust: WHO | 1.19 ***<br>[1.13 – 1.24] | 1.10 **<br>[1.03 – 1.18] | 0.96<br>[0.71 – 1.28] | 1.57 **<br>[1.15 – 2.14] | 1.00<br>[0.74 – 1.33] | 1.03<br>[0.78 – 1.35] | 1.11<br>[0.74 – 1.63] | 0.90<br>[0.69 – 1.18] | 1.07<br>[0.83 – 1.38] | 1.12<br>[0.82 – 1.53] | 1.59 ***<br>[1.24 – 2.04] | 1.15<br>[0.91 – 1.45] | 1.28 *<br>[1.02 – 1.60] |
| Personal efficacy | 1.01<br>[0.97 – 1.06] | 0.93 *<br>[0.87 – 0.99] | 0.85<br>[0.64 – 1.12] | 1.05<br>[0.81 – 1.35] | 0.93<br>[0.71 – 1.21] | 0.91<br>[0.71 – 1.16] | 1.02<br>[0.73 – 1.43] | 0.92<br>[0.72 – 1.18] | 1.01<br>[0.75 – 1.36] | 0.86<br>[0.62 – 1.17] | 0.82<br>[0.64 – 1.05] | 0.74 **<br>[0.60 – 0.92] | 1.09<br>[0.90 – 1.33] |
| Govt efficacy | 1.01<br>[0.96 – 1.07] | 1.07<br>[0.98 – 1.16] | 1.25<br>[0.88 – 1.79] | 0.94<br>[0.70 – 1.26] | 0.97<br>[0.70 – 1.33] | 1.08<br>[0.79 – 1.47] | 1.35<br>[0.92 – 1.98] | 1.15<br>[0.81 – 1.64] | 1.05<br>[0.75 – 1.46] | 1.00<br>[0.68 – 1.50] | 0.89<br>[0.66 – 1.19] | 1.20<br>[0.89 – 1.61] | 1.13<br>[0.85 – 1.50] |
| Perceived infection risk | 1.12 ***<br>[1.07 – 1.16] | 1.13 ***<br>[1.07 – 1.20] | 1.06<br>[0.82 – 1.37] | 1.49 **<br>[1.16 – 1.92] | 1.00<br>[0.80 – 1.25] | 1.11<br>[0.89 – 1.38] | 1.47 **<br>[1.11 – 1.96] | 1.20<br>[0.97 – 1.48] | 1.07<br>[0.83 – 1.38] | 1.43 *<br>[1.06 – 1.92] | 1.27 *<br>[1.02 – 1.57] | 1.30 **<br>[1.07 – 1.58] | 0.79 *<br>[0.65 – 0.97] |
| Worry about COVID | 1.49 ***<br>[1.43 – 1.55] | 1.53 ***<br>[1.44 – 1.62] | 1.69 ***<br>[1.34 – 2.16] | 1.34 *<br>[1.05 – 1.70] | 1.37 *<br>[1.07 – 1.74] | 1.19<br>[0.95 – 1.48] | 1.55 **<br>[1.18 – 2.03] | 1.33 **<br>[1.07 – 1.65] | 1.24<br>[0.96 – 1.59] | 1.66 ***<br>[1.26 – 2.21] | 1.31 *<br>[1.06 – 1.63] | 1.37 **<br>[1.13 – 1.66] | 1.77 ***<br>[1.44 – 2.19] |
| Observations | 19256 | 8418 | 644 | 641 | 669 | 666 | 532 | 590 | 677 | 629 | 684 | 653 | 656 |
| R<sup>2</sup> Tjur | 0.126 | 0.102 | 0.096 | 0.183 | 0.061 | 0.137 | 0.142 | 0.077 | 0.04 | 0.115 | 0.196 | 0.102 | 0.137 |

Odd ratios [95CI] shown, all continuous measure were standardized (scaled and mean-centered) prior to analysis. For space, samples are defined by their two character ISO country code and a letter denoting participant source (D, Dynata; R, Respondi; P, Prolific). <sup>a</sup>Gender is unstandardized
\*p < .05, \*\*p < .01, \*\*\* p < .001

**Table S6.**
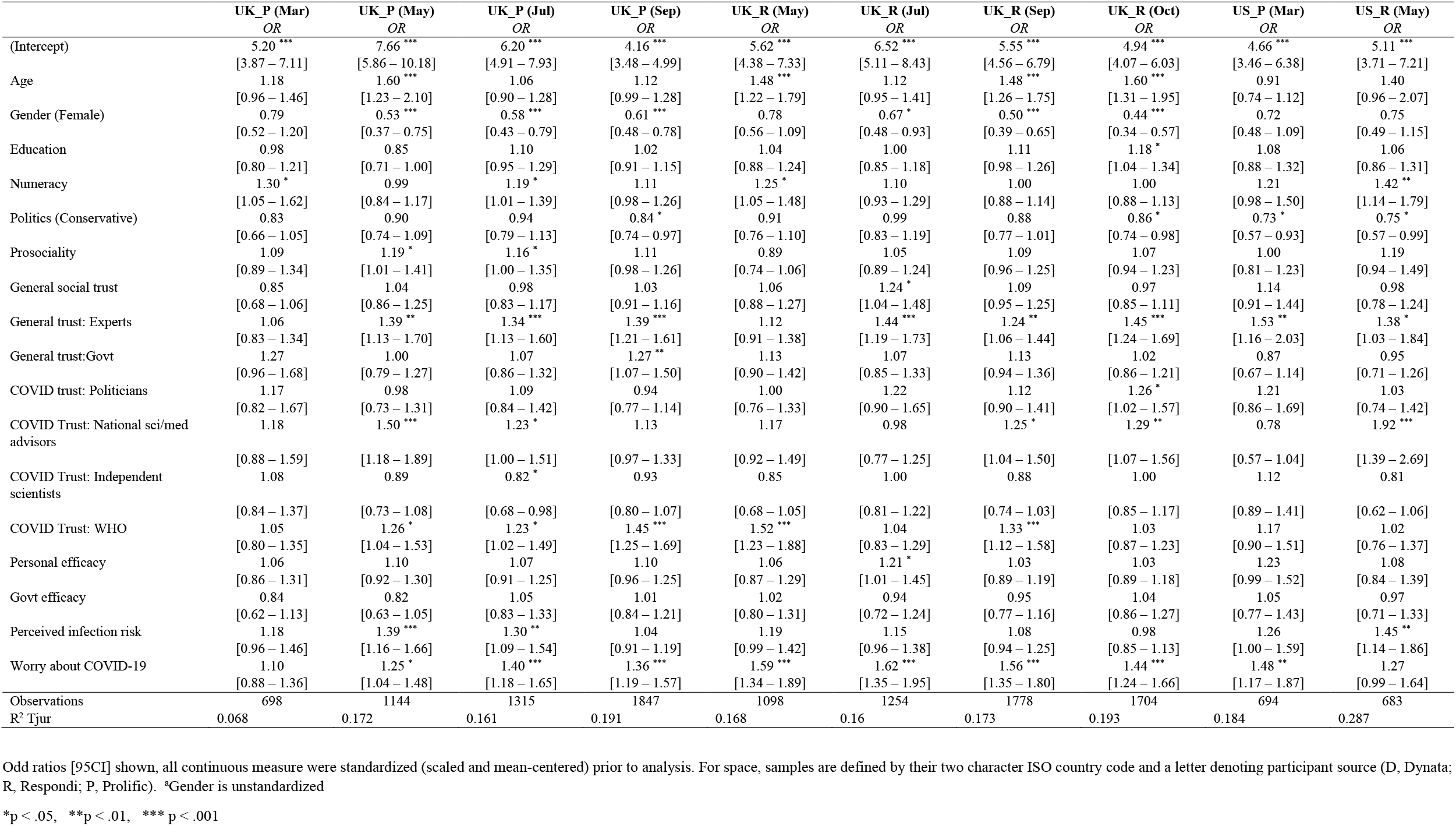
Full logistic regression results from model predicting vaccine acceptance, UK and US samples

**Figure S1.**
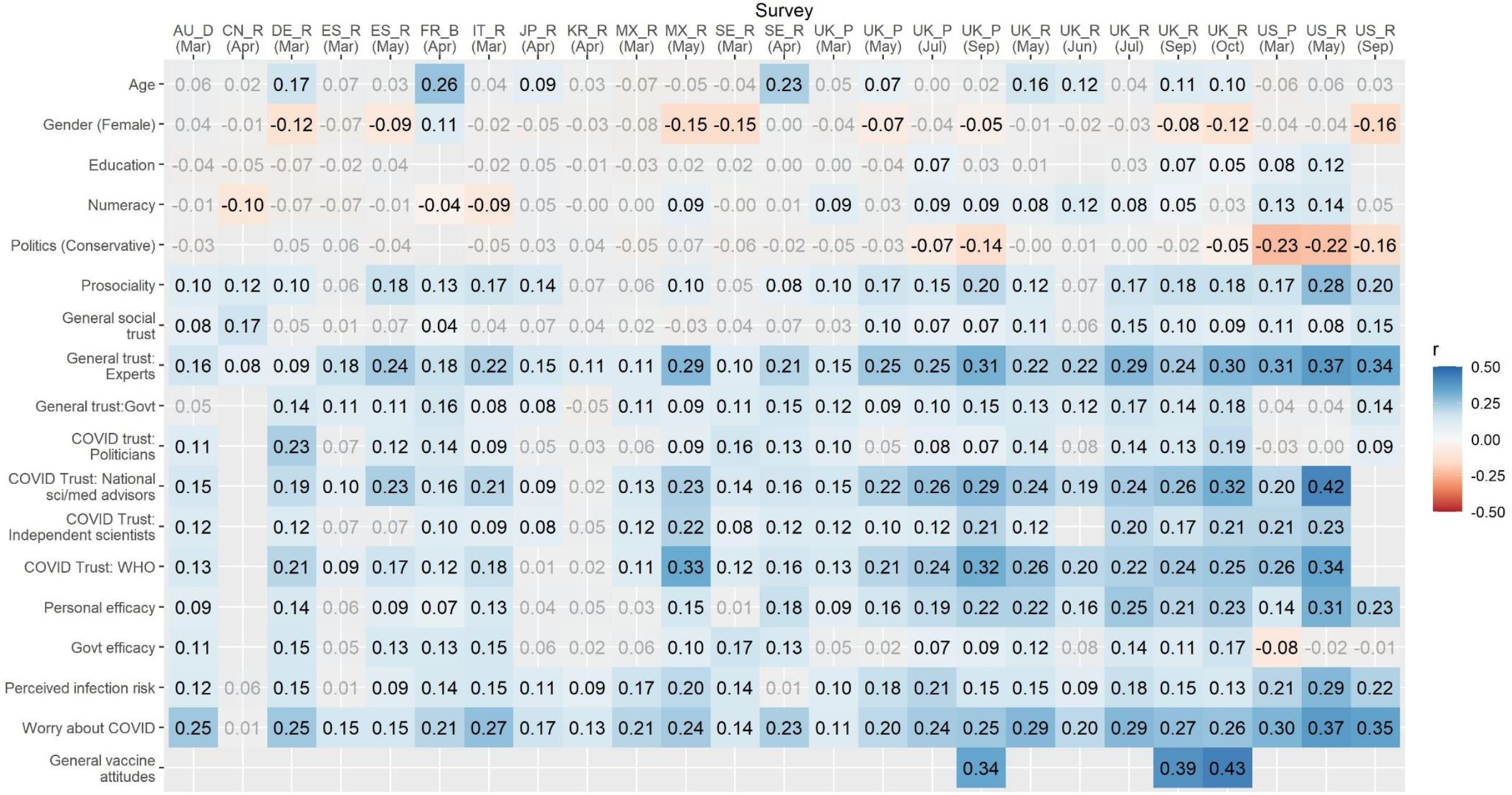
Point biserial correlations between predictors and vaccine acceptance across all samples. Greyed values are non-significant (*p* > .05). Blank spaces indicate predictors which were not included in a given survey.

**Figure S2.**
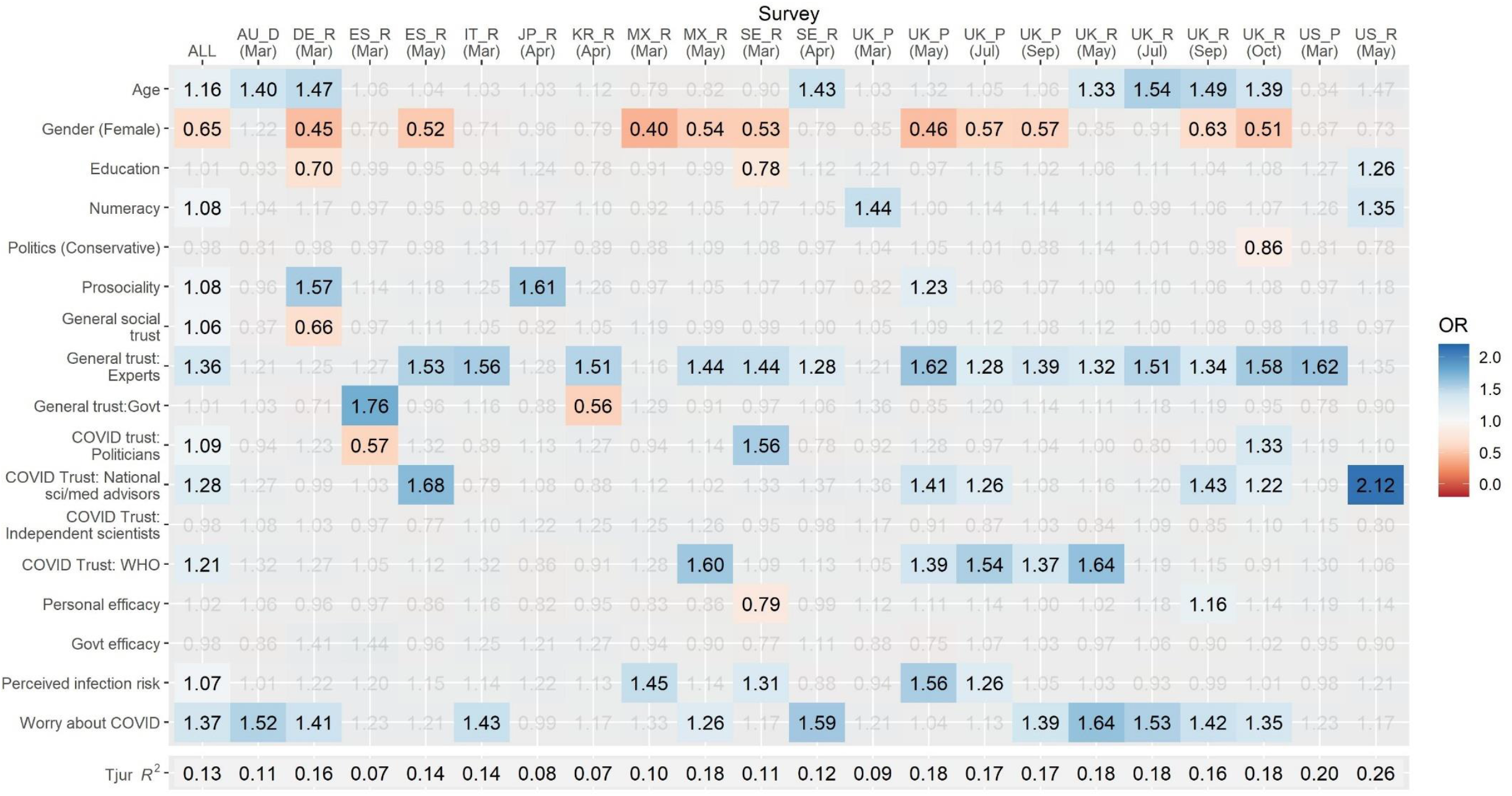
Heatmap of odds ratios in model predicting recommending vaccine to vulnerable friends/family. Columns represent individual samples and rows represent predictors in model. Grey values are non-significant, p > .05. Red shading indicates a lower likelihood of vaccine acceptance and blue shading a higher likelihood. For space, samples are defined by their two character ISO country code and a letter denoting participant source (D, Dynata; R, Respondi; P, Prolific).

**Table S7.**
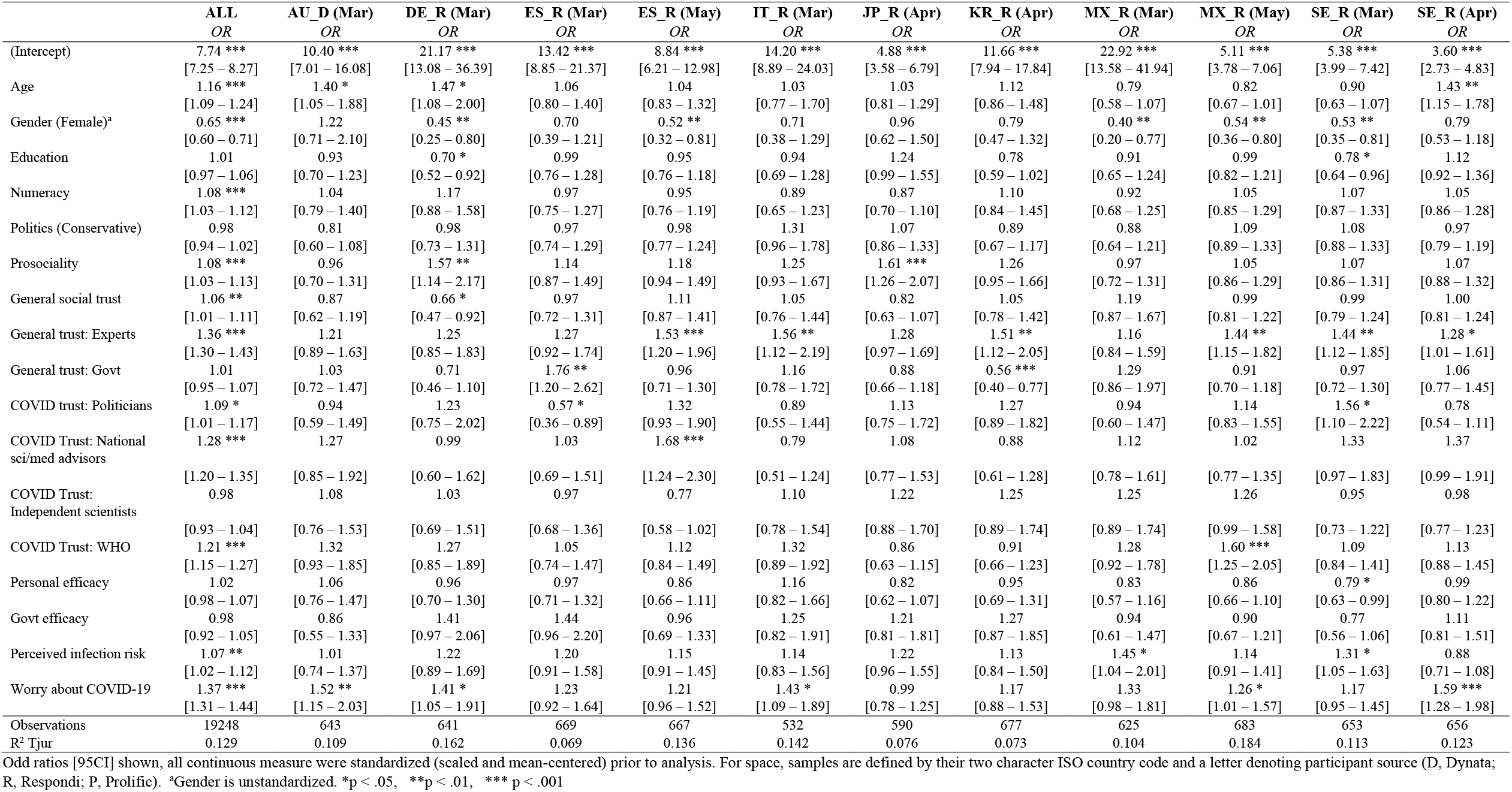
Full logistic regression results from model predicting vaccine recommendation to vulnerable others, excluding UK and US samples (shown in Table S8)

|  | ALL<br>OR | AU_D (Mar)<br>OR | DE_R (Mar)<br>OR | ES_R (Mar)<br>OR | ES_R (May)<br>OR | IT_R (Mar)<br>OR | JP_R (Apr)<br>OR | KR_R (Apr)<br>OR | MX_R (Mar)<br>OR | MX_R (May)<br>OR | SE_R (Mar)<br>OR | SE_R (Apr)<br>OR |
| --- | --- | --- | --- | --- | --- | --- | --- | --- | --- | --- | --- | --- |
| (Intercept) | 7.74 ***<br>[7.25 – 8.27] | 10.40 ***<br>[7.01 – 16.08] | 21.17 ***<br>[13.08 – 36.39] | 13.42 ***<br>[8.85 – 21.37] | 8.84 ***<br>[6.21 – 12.98] | 14.20 ***<br>[8.89 – 24.03] | 4.88 ***<br>[3.58 – 6.79] | 11.66 ***<br>[7.94 – 17.84] | 22.92 ***<br>[13.58 – 41.94] | 5.11 ***<br>[3.78 – 7.06] | 5.38 ***<br>[3.99 – 7.42] | 3.60 ***<br>[2.73 – 4.83] |
| Age | 1.16 ***<br>[1.09 – 1.24] | 1.40 *<br>[1.05 – 1.88] | 1.47 *<br>[1.08 – 2.00] | 1.06<br>[0.80 – 1.40] | 1.04<br>[0.83 – 1.32] | 1.03<br>[0.77 – 1.70] | 1.03<br>[0.81 – 1.29] | 1.12<br>[0.86 – 1.48] | 0.79<br>[0.58 – 1.07] | 0.82<br>[0.67 – 1.01] | 0.90<br>[0.63 – 1.07] | 1.43 **<br>[1.15 – 1.78] |
| Gender (Female)<sup>a</sup> | 0.65 ***<br>[0.60 – 0.71] | 1.22<br>[0.71 – 2.10] | 0.45 **<br>[0.25 – 0.80] | 0.70<br>[0.39 – 1.21] | 0.52 **<br>[0.32 – 0.81] | 0.71<br>[0.38 – 1.29] | 0.96<br>[0.62 – 1.50] | 0.79<br>[0.47 – 1.32] | 0.40 **<br>[0.20 – 0.77] | 0.54 **<br>[0.36 – 0.80] | 0.53 **<br>[0.35 – 0.81] | 0.79<br>[0.53 – 1.18] |
| Education | 1.01<br>[0.97 – 1.06] | 0.93<br>[0.70 – 1.23] | 0.70 *<br>[0.52 – 0.92] | 0.99<br>[0.76 – 1.28] | 0.95<br>[0.76 – 1.18] | 0.94<br>[0.69 – 1.28] | 1.24<br>[0.99 – 1.55] | 0.78<br>[0.59 – 1.02] | 0.91<br>[0.65 – 1.24] | 0.99<br>[0.82 – 1.21] | 0.78 *<br>[0.64 – 0.96] | 1.12<br>[0.92 – 1.36] |
| Numeracy | 1.08 ***<br>[1.03 – 1.12] | 1.04<br>[0.79 – 1.40] | 1.17<br>[0.88 – 1.58] | 0.97<br>[0.75 – 1.27] | 0.95<br>[0.76 – 1.19] | 0.89<br>[0.65 – 1.23] | 0.87<br>[0.70 – 1.10] | 1.10<br>[0.84 – 1.45] | 0.92<br>[0.68 – 1.25] | 1.05<br>[0.85 – 1.29] | 1.07<br>[0.87 – 1.33] | 1.05<br>[0.86 – 1.28] |
| Politics (Conservative) | 0.98<br>[0.94 – 1.02] | 0.81<br>[0.60 – 1.08] | 0.98<br>[0.73 – 1.31] | 0.97<br>[0.74 – 1.29] | 0.98<br>[0.77 – 1.24] | 1.31<br>[0.96 – 1.78] | 1.07<br>[0.86 – 1.33] | 0.89<br>[0.67 – 1.17] | 0.88<br>[0.64 – 1.21] | 1.09<br>[0.89 – 1.33] | 1.08<br>[0.88 – 1.33] | 0.97<br>[0.79 – 1.19] |
| Prosociality | 1.08 ***<br>[1.03 – 1.13] | 0.96<br>[0.70 – 1.31] | 1.57 **<br>[1.14 – 2.17] | 1.14<br>[0.87 – 1.49] | 1.18<br>[0.94 – 1.49] | 1.25<br>[0.93 – 1.67] | 1.61 ***<br>[1.26 – 2.07] | 1.26<br>[0.95 – 1.66] | 0.97<br>[0.72 – 1.31] | 1.05<br>[0.86 – 1.29] | 1.07<br>[0.86 – 1.31] | 1.07<br>[0.88 – 1.32] |
| General social trust | 1.06 **<br>[1.01 – 1.11] | 0.87<br>[0.62 – 1.19] | 0.66 *<br>[0.47 – 0.92] | 0.97<br>[0.72 – 1.31] | 1.11<br>[0.87 – 1.41] | 1.05<br>[0.76 – 1.44] | 0.82<br>[0.63 – 1.07] | 1.05<br>[0.78 – 1.42] | 1.19<br>[0.87 – 1.67] | 0.99<br>[0.81 – 1.22] | 0.99<br>[0.79 – 1.24] | 1.00<br>[0.81 – 1.24] |
| General trust: Experts | 1.36 ***<br>[1.30 – 1.43] | 1.21<br>[0.89 – 1.63] | 1.25<br>[0.85 – 1.83] | 1.27<br>[0.92 – 1.74] | 1.53 ***<br>[1.20 – 1.96] | 1.56 **<br>[1.12 – 2.19] | 1.28<br>[0.97 – 1.69] | 1.51 **<br>[1.12 – 2.05] | 1.16<br>[0.84 – 1.59] | 1.44 **<br>[1.15 – 1.82] | 1.44 **<br>[1.12 – 1.85] | 1.28 *<br>[1.01 – 1.61] |
| General trust: Govt | 1.01<br>[0.95 – 1.07] | 1.03<br>[0.72 – 1.47] | 0.71<br>[0.46 – 1.10] | 1.76 **<br>[1.20 – 2.62] | 0.96<br>[0.71 – 1.30] | 1.16<br>[0.78 – 1.72] | 0.88<br>[0.66 – 1.18] | 0.56 ***<br>[0.40 – 0.77] | 1.29<br>[0.86 – 1.97] | 0.91<br>[0.70 – 1.18] | 0.97<br>[0.72 – 1.30] | 1.06<br>[0.77 – 1.45] |
| COVID trust: Politicians | 1.09 *<br>[1.01 – 1.17] | 0.94<br>[0.59 – 1.49] | 1.23<br>[0.75 – 2.02] | 0.57 *<br>[0.36 – 0.89] | 1.32<br>[0.93 – 1.90] | 0.89<br>[0.55 – 1.44] | 1.13<br>[0.75 – 1.72] | 1.27<br>[0.89 – 1.82] | 0.94<br>[0.60 – 1.47] | 1.14<br>[0.83 – 1.55] | 1.56 *<br>[1.10 – 2.22] | 0.78<br>[0.54 – 1.11] |
| COVID Trust: National<br>sci/med advisors | 1.28 ***<br>[1.20 – 1.35] | 1.27<br>[0.85 – 1.92] | 0.99<br>[0.60 – 1.62] | 1.03<br>[0.69 – 1.51] | 1.68 ***<br>[1.24 – 2.30] | 0.79<br>[0.51 – 1.24] | 1.08<br>[0.77 – 1.53] | 0.88<br>[0.61 – 1.28] | 1.12<br>[0.78 – 1.61] | 1.02<br>[0.77 – 1.35] | 1.33<br>[0.97 – 1.83] | 1.37<br>[0.99 – 1.91] |
| COVID Trust:<br>Independent scientists | 0.98<br>[0.93 – 1.04] | 1.08<br>[0.76 – 1.53] | 1.03<br>[0.69 – 1.51] | 0.97<br>[0.68 – 1.36] | 0.77<br>[0.58 – 1.02] | 1.10<br>[0.78 – 1.54] | 1.22<br>[0.88 – 1.70] | 1.25<br>[0.89 – 1.74] | 1.25<br>[0.89 – 1.74] | 1.26<br>[0.99 – 1.58] | 0.95<br>[0.73 – 1.22] | 0.98<br>[0.77 – 1.23] |
| COVID Trust: WHO | 1.21 ***<br>[1.15 – 1.27] | 1.32<br>[0.93 – 1.85] | 1.27<br>[0.85 – 1.89] | 1.05<br>[0.74 – 1.47] | 1.12<br>[0.84 – 1.49] | 1.32<br>[0.89 – 1.92] | 0.86<br>[0.63 – 1.15] | 0.91<br>[0.66 – 1.23] | 1.28<br>[0.92 – 1.78] | 1.60 ***<br>[1.25 – 2.05] | 1.09<br>[0.84 – 1.41] | 1.13<br>[0.88 – 1.45] |
| Personal efficacy | 1.02<br>[0.98 – 1.07] | 1.06<br>[0.76 – 1.47] | 0.96<br>[0.70 – 1.30] | 0.97<br>[0.71 – 1.32] | 0.86<br>[0.66 – 1.11] | 1.16<br>[0.82 – 1.66] | 0.82<br>[0.62 – 1.07] | 0.95<br>[0.69 – 1.31] | 0.83<br>[0.57 – 1.16] | 0.86<br>[0.66 – 1.10] | 0.79 *<br>[0.63 – 0.99] | 0.99<br>[0.80 – 1.22] |
| Govt efficacy | 0.98<br>[0.92 – 1.05] | 0.86<br>[0.55 – 1.33] | 1.41<br>[0.97 – 2.06] | 1.44<br>[0.96 – 2.20] | 0.96<br>[0.69 – 1.33] | 1.25<br>[0.82 – 1.91] | 1.21<br>[0.81 – 1.81] | 1.27<br>[0.87 – 1.85] | 0.94<br>[0.61 – 1.47] | 0.90<br>[0.67 – 1.21] | 0.77<br>[0.56 – 1.06] | 1.11<br>[0.81 – 1.51] |
| Perceived infection risk | 1.07 **<br>[1.02 – 1.12] | 1.01<br>[0.74 – 1.37] | 1.22<br>[0.89 – 1.69] | 1.20<br>[0.91 – 1.58] | 1.15<br>[0.91 – 1.45] | 1.14<br>[0.83 – 1.56] | 1.22<br>[0.96 – 1.55] | 1.13<br>[0.84 – 1.50] | 1.45 *<br>[1.04 – 2.01] | 1.14<br>[0.91 – 1.41] | 1.31 *<br>[1.05 – 1.63] | 0.88<br>[0.71 – 1.08] |
| Worry about COVID-19 | 1.37 ***<br>[1.31 – 1.44] | 1.52 **<br>[1.15 – 2.03] | 1.41 *<br>[1.05 – 1.91] | 1.23<br>[0.92 – 1.64] | 1.21<br>[0.96 – 1.52] | 1.43 *<br>[1.09 – 1.89] | 0.99<br>[0.78 – 1.25] | 1.17<br>[0.88 – 1.53] | 1.33<br>[0.98 – 1.81] | 1.26 *<br>[1.01 – 1.57] | 1.17<br>[0.95 – 1.45] | 1.59 ***<br>[1.28 – 1.98] |
| Observations | 19248 | 643 | 641 | 669 | 667 | 532 | 590 | 677 | 625 | 683 | 653 | 656 |
| R<sup>2</sup> Tjur | 0.129 | 0.109 | 0.162 | 0.069 | 0.136 | 0.142 | 0.076 | 0.073 | 0.104 | 0.184 | 0.113 | 0.123 |

Odd ratios [95CI] shown, all continuous measure were standardized (scaled and mean-centered) prior to analysis. For space, samples are defined by their two character ISO country code and a letter denoting participant source (D, Dynata; R, Respondi; P, Prolific). <sup>a</sup>Gender is unstandardized. \*p < .05, \*\*p < .01, \*\*\* p < .001

**Table S8.**
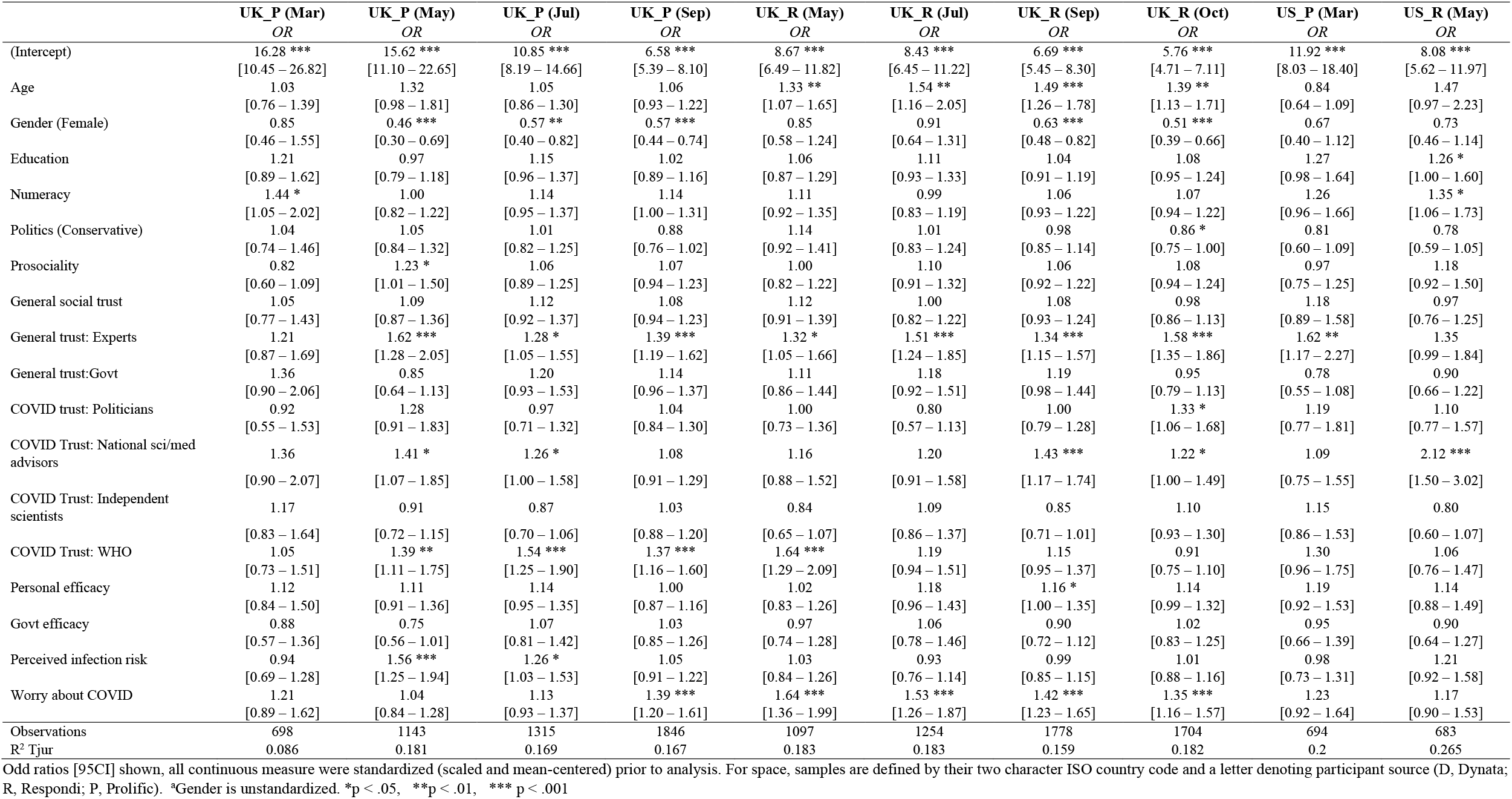
Full logistic regression results from model predicting vaccine recommendation to vulnerable others, UK and US samples

**Table S9.**
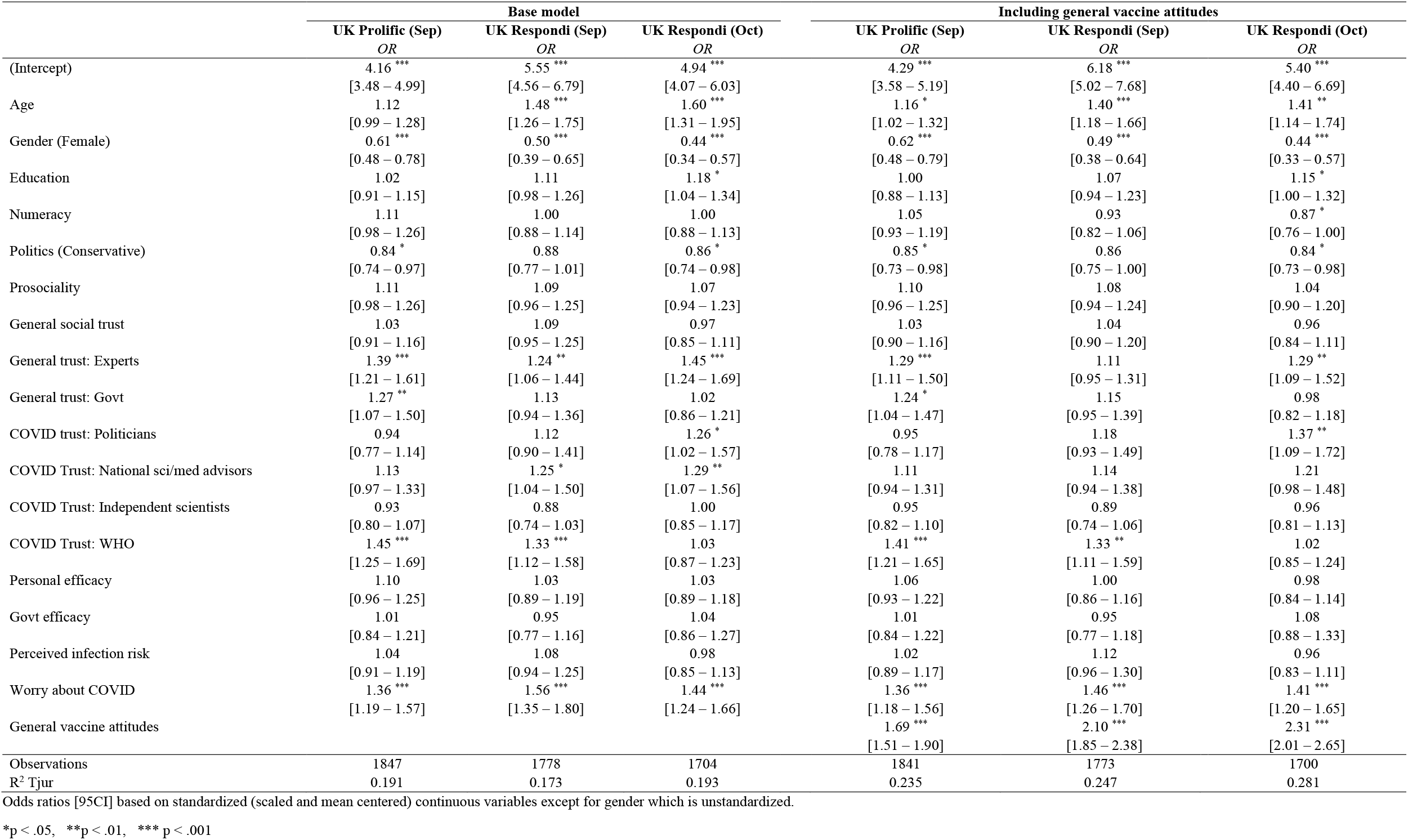
Result of logistic regression models predicting vaccine acceptance, including or excluding general vaccine attitudes.

## Footnotes

1 Based on respondents who answered the question. In the Italy sample a number of participants were not presented with these items due to a technical error (*n* = 80, 11%). In the remaining samples the average proportion of missing responses for vaccine intention and recommendation items was 1%.

2 UK data was over represented in our pooled sample. As a robustness check we also fitted the model to the pooled sample with UK data removed and report that the effects of gender, trust in experts and worry remain significant (*p*s < .001; see Table S5).

